# Blood transcriptomic signatures predict poor treatment outcomes in drug-susceptible pulmonary TB in Brazil

**DOI:** 10.1101/2025.10.07.25337480

**Authors:** Simon C Mendelsohn, Bruno B Andrade, Mariana Araújo-Pereira, Gustavo C Amorim, Mzwandile Erasmus, Alice MS Andrade, Michelle Fisher, Marina C Figueiredo, Vanessa M Muwanga, Afranio L Kritski, Marcelo Cordeiro-Santos, Valeria C Rolla, Mark Hatherill, Timothy R Sterling, Thomas J Scriba, the RePORT South Africa, RePORT-Brazil Consortia

## Abstract

**Background:** Non-sputum biomarkers to monitor tuberculosis (TB) treatment and predict poor outcomes are lacking. We evaluated host-blood transcriptomic signatures for treatment monitoring and prognosis (death, treatment failure, recurrence) in adults with pulmonary TB.

**Methods:** Adults with culture-confirmed, drug-susceptible pulmonary TB were enrolled at five Brazilian sites. Whole-blood PAXgene samples were collected at baseline, month 2 (M2), and end of treatment (EoT). Treatment failure was defined as sputum culture positivity at month 5 or later. Participants were followed for 24 months from treatment initiation for clinical or microbiological TB recurrence. Unfavourable outcomes were matched ∼1:3 to recurrence-free cure. Twenty-two published blood transcriptomic signatures were measured by microfluidic RT-qPCR and benchmarked against the WHO Target Product Profile (TPP) criteria.

**Results:** We matched 263 participants with recurrence-free cure to 33 with treatment failure, 24 who died (TB/unknown cause), and 9 with recurrence. Signature scores generally declined from baseline to EoT. Multiple signatures predicted recurrence at baseline and M2 (AUC range 0.71-0.91), with waning performance at EoT (AUC range 0.42-0.89). Against the WHO TPP, 2/22 signatures met minimum criteria at baseline, 13/22 at M2, and none at EoT. Prediction of treatment failure was poor across timepoints (AUC <0.70). In contrast, Thompson5 and others at baseline predicted death during treatment or follow-up (AUC ≥0.80).

**Conclusion:** Blood transcriptomic signatures tracked treatment response and predicted recurrence and death, meeting WHO TPP benchmarks at baseline and M2. These findings support prospective, biomarker-guided trials to individualise TB therapy—shortening regimens for early responders and intensifying care for high-risk patients.

**Funding:** This work was supported by the U.S. National Institutes of Health, CRDF Global, and Departamento de Ciência e Tecnologia (DECIT) - Secretaria de Ciência e Tecnologia (SCTIE), Ministério da Saúde (MS), Brazil.

## INTRODUCTION

Despite effective antibiotics, tuberculosis (TB) treatment failure, recurrence, and death remain major challenges, especially in high-risk populations. In 2021-2022, global treatment failure rates were estimated at 12% for new and relapse TB cases, increasing to 21% among people living with HIV (PLHIV), 22% in previously treated individuals (excluding relapse), and 32% among those with multidrug- or rifampicin-resistant TB.^1^ World Health Organization (WHO) guidelines for drug-susceptible pulmonary TB recommend sputum smear microscopy or culture at the end of the intensive phase to monitor treatment response.^2^ However, both have significant limitations.^3^ Culture is slow, and smear microscopy cannot distinguish viable from dead bacilli, or *Mycobacterium tuberculosis* (Mtb) from nontuberculous mycobacteria. Both methods rely on sputum, which is often unavailable—particularly in children, PLHIV, those with good response to treatment, and those with extrapulmonary or paucibacillary disease.^4^ Nucleic acid amplification tests, such as Xpert MTB/RIF, may remain positive despite effective therapy due to persistence of DNA from dead mycobacteria. Consequently, treatment response is often assessed by non-specific and insensitive clinical measures, such as weight gain and symptom improvement. No validated non-sputum biomarkers currently exist in clinical practice to guide TB treatment decisions.

Standard regimens for drug-susceptible pulmonary TB, either the 6-month (2HRZE/4HR), or more recently, the 4-month rifapentine-moxifloxacin-containing regimen, are prescribed irrespective of disease severity.^2^ Some expert consensus guidelines recommend extending to a total of 9 months (2HRZE/7HR) in patients with cavitation on the initial chest radiograph and a positive culture at 2 months.^5^ These regimens include four potentially toxic drugs, which can cause adverse effects and contribute to poor adherence.^6^ However, clinical trials have shown that cure is possible in selected individuals with shorter regimens.^7,8^ To enable individualized treatment duration, rapid and accessible biomarkers—ideally from non-sputum samples like finger-prick blood—are needed to predict early cure or failure before treatment completion.

Peripheral blood transcriptomic signatures offer a promising non-sputum approach. Several host-response blood signatures have been associated with TB treatment response,^9–16^ including RISK6, which correlates with lung inflammation on PET-CT and inversely with sputum mycobacterial load.^13^ The prognostic Zak16 transcriptomic signature tracked disease resolution and identified patients at risk of failure within four weeks of starting treatment.^12^ These findings have been replicated in HIV-negative and HIV-positive populations across South Africa, Brazil, and a multi-country study in Bangladesh, Georgia, Lebanon, and Madagascar.^13,17^ In comparative studies, transcriptomic signatures outperformed traditional biomarkers like C-reactive protein.^18^ In the Catalysis cohort, RISK6, Zak16, and Sweeney3 signatures distinguished TB patients with cure from those with treatment failure as early as diagnosis, and with high accuracy at treatment end.^12–14^ Despite promising results, most studies validating transcriptomic signatures have been limited by small sample sizes and few adverse outcomes. To our knowledge, no large prospective cohort has evaluated the performance of multiple RT-qPCR-based transcriptomic signatures for predicting TB treatment failure, recurrence, or death.

Accurate, non-sputum biomarkers are critically needed to monitor TB treatment response and guide therapy modification. Transcriptomic signatures could support regimen shortening in early responders, intensification for slow responders, and continuation for those at risk of relapse. Such signatures may also be used to expedite drug development and treatment trials.^18^ Host-blood transcriptomic signatures are now feasible for point-of-care implementation.^19–22^ We evaluated parsimonious host-response blood transcriptomic signatures to monitor TB treatment and predict adverse outcomes in adults with drug-susceptible pulmonary TB.

## METHODS

### Study design and participants

Regional Prospective Observational Research for Tuberculosis (RePORT)-Brazil is a prospective observational cohort study that enrolled participants at five centres: three in the state of Rio de Janeiro (City of Rio de Janeiro: [1] Instituto Nacional de Infectologia and [2] Clínica da Família Rinaldo Delamare. Duque de Caxias: [3] Duque de Caxias Health Center), one in Salvador (Instituto Brasileiro para Investigação da Tuberculose), and one in Manaus (Fundação Medicina Tropical Dr. Heitor Vieira Dourado). Details of the study cohort have been published previously.^23^ This was a case-control study nested within the observational cohort. For this analysis, adults (≥18 years) initiating treatment for culture-confirmed drug-susceptible pulmonary TB were included. Sputum was solicited at month 5 of treatment for culture, and TB treatment failure was defined as a culture positive for Mtb. Cause of death during treatment or study follow-up was adjudicated by site investigators and classified as: (i) deaths directly attributable to TB; (ii) deaths from unknown causes (possibly related to TB or with TB as a contributing factor); and (iii) deaths from unrelated causes (e.g., trauma, non-natural causes). Deaths from unrelated causes were excluded from analyses. TB recurrence was evaluated over 24 months following treatment initiation through symptom-triggered investigations (e.g., ≥2 weeks of cough, fever, night sweats, or weight loss, or any haemoptysis), which included chest radiography and sputum smear and culture testing. Microbiologically-confirmed recurrence required at least one sputum specimen culture-positive for Mtb. Clinical recurrence, determined by site investigators, was defined by compatible signs and symptoms of TB, with or without histopathology suggestive of TB (e.g., necrotizing or caseating granulomas). Individuals with treatment failure or recurrent TB were treated or re-treated according to local guidelines.

### Blood sample collection and RNA extraction

Venous blood (2.5mL) was collected in PAXgene RNA tubes (PreAnalytiX, Switzerland) at baseline, month 2, and at the end of TB treatment (EoT) for all participants, frozen at −20°C, and shipped to the Instituto Gonçalo Moniz, Fundação Oswaldo Cruz in Salvador, Brazil. Additional PAXgene samples were collected at recurrence visits in participants with TB recurrence. Samples were thawed and RNA was manually extracted using the PAXgene Blood RNA Kit (Qiagen, Germany) according to the manufacturer’s instructions. Approximately 50ng of RNA (∼8.3ng/µL in 6µL water) per sample were plated into 96-well PCR plates, frozen at −80°C, and shipped to the South African Tuberculosis Vaccine Initiative (SATVI) in Cape Town, South Africa.

### Sample size and matching

All available RNA samples from individuals with unfavourable outcomes (treatment failure, death, or TB recurrence) were matched to participants who completed follow-up with recurrence-free cure. Participants who were lost to follow-up or withdrawn were excluded from the matching procedure. Propensity (or risk) scores^24^ were estimated using logistic regression (**Supplementary Methods**).

### Transcriptomic signature panel design and measurement

Published TB transcriptomic signatures were selected based on diagnostic and prognostic performance reported in recent systematic reviews and head-to-head comparisons,^25–29^ as well as ongoing work at the SATVI laboratory.^30^ Most of the included signatures map to interferon (IFN) response and inflammatory gene pathways. We included 22 parsimonious signatures—defined as those derived using feature reduction during discovery—based on: (i) number of transcripts (<30); (ii) availability of primer-probe sequences or predesigned TaqMan assays; and (iii) evaluation in at least one external validation cohort (**Table 1**). Signatures were translated to a microfluidic, multiplex real-time quantitative PCR (RT-qPCR) platform, reparameterised, and measured as previously described (**Supplementary Methods**), using a panel of pre-qualified TaqMan gene expression primer-probe assays (**Table S1**).^30–32^

**Table 1.**
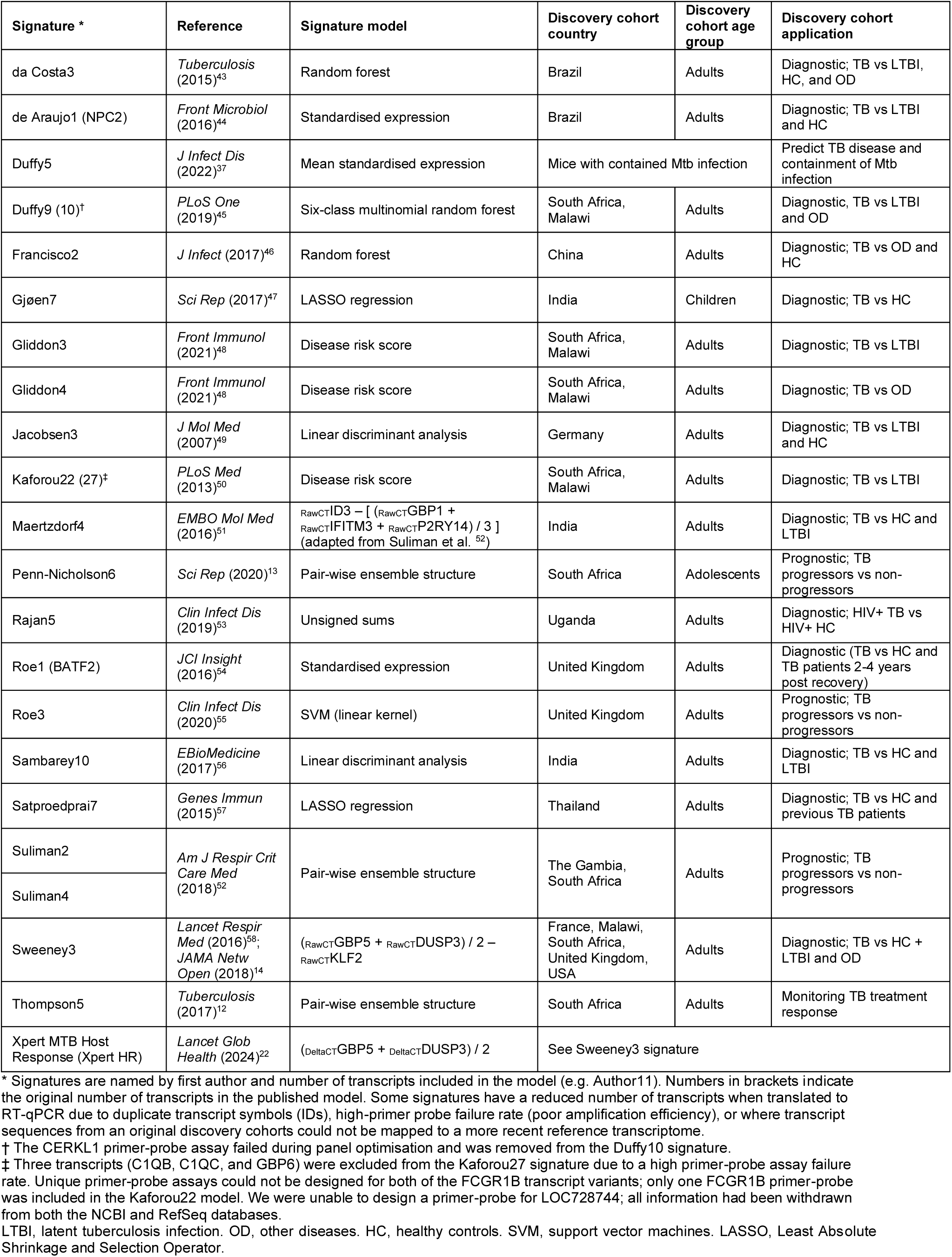
Characteristics of host blood transcriptomic signatures included in the analysis.

| Signature * | Reference | Signature model | Discovery cohort country | Discovery cohort age group | Discovery cohort application |
| --- | --- | --- | --- | --- | --- |
| da Costa3 | *Tuberculosis* (2015)<sup>43</sup> | Random forest | Brazil | Adults | Diagnostic; TB vs LTBI, HC, and OD |
| de Araujo1 (NPC2) | *Front Microbiol* (2016)<sup>44</sup> | Standardised expression | Brazil | Adults | Diagnostic; TB vs LTBI and HC |
| Duffy5 | *J Infect Dis* (2022)<sup>37</sup> | Mean standardised expression | Mice with contained Mtb infection |  | Predict TB disease and containment of Mtb infection |
| Duffy9 (10)<sup>†</sup> | *PLoS One* (2019)<sup>45</sup> | Six-class multinomial random forest | South Africa, Malawi | Adults | Diagnostic; TB vs LTBI and OD |
| Francisco2 | *J Infect* (2017)<sup>46</sup> | Random forest | China | Adults | Diagnostic; TB vs OD and HC |
| GjØen7 | *Sci Rep* (2017)<sup>47</sup> | LASSO regression | India | Children | Diagnostic; TB vs HC |
| Gliddon3 | *Front Immunol* (2021)<sup>48</sup> | Disease risk score | South Africa, Malawi | Adults | Diagnostic; TB vs LTBI |
| Gliddon4 | *Front Immunol* (2021)<sup>48</sup> | Disease risk score | South Africa, Malawi | Adults | Diagnostic; TB vs OD |
| Jacobsen3 | *J Mol Med* (2007)<sup>49</sup> | Linear discriminant analysis | Germany | Adults | Diagnostic; TB vs LTBI and HC |
| Kaforou22 (27)<sup>‡</sup> | *PLoS Med* (2013)<sup>50</sup> | Disease risk score | South Africa, Malawi | Adults | Diagnostic; TB vs LTBI |
| Maertzdorf4 | *EMBO Mol Med* (2016)<sup>51</sup> | $\text{RawCTID3} - [(\text{RawCTGBP1} + \text{RawCTIFITM3} + \text{RawCTP2RY14}) / 3]$ (adapted from Suliman et al. <sup>52</sup>) | India | Adults | Diagnostic; TB vs HC and LTBI |
| Penn-Nicholson6 | *Sci Rep* (2020)<sup>13</sup> | Pair-wise ensemble structure | South Africa | Adolescents | Prognostic; TB progressors vs non-progressors |
| Rajan5 | *Clin Infect Dis* (2019)<sup>53</sup> | Unsigned sums | Uganda | Adults | Diagnostic; HIV+ TB vs HIV+ HC |
| Roe1 (BATF2) | *JCI Insight* (2016)<sup>54</sup> | Standardised expression | United Kingdom | Adults | Diagnostic (TB vs HC and TB patients 2-4 years post recovery) |
| Roe3 | *Clin Infect Dis* (2020)<sup>55</sup> | SVM (linear kernel) | United Kingdom | Adults | Prognostic; TB progressors vs non-progressors |
| Sambarey10 | *EBioMedicine* (2017)<sup>56</sup> | Linear discriminant analysis | India | Adults | Diagnostic; TB vs HC and LTBI |
| Satproedprai7 | *Genes Immun* (2015)<sup>57</sup> | LASSO regression | Thailand | Adults | Diagnostic; TB vs HC and previous TB patients |
| Suliman2 | *Am J Respir Crit Care Med* (2018)<sup>52</sup> | Pair-wise ensemble structure | The Gambia, South Africa | Adults | Prognostic; TB progressors vs non-progressors |
| Suliman4 |  |  |  |  |  |
| Sweeney3 | *Lancet Respir Med* (2016)<sup>58</sup>, *JAMA Netw Open* (2018)<sup>14</sup> | $(\text{RawCTGBP5} + \text{RawCTDUSP3}) / 2 - \text{RawCTKLF2}$ | France, Malawi, South Africa, United Kingdom, USA | Adults | Diagnostic; TB vs HC + LTBI and OD |
| Thompson5 | *Tuberculosis* (2017)<sup>12</sup> | Pair-wise ensemble structure | South Africa | Adults | Monitoring TB treatment response |
| Xpert MTB Host Response (Xpert HR) | *Lancet Glob Health* (2024)<sup>22</sup> | $(\text{DeltaCTGBP5} + \text{DeltaCTDUSP3}) / 2$ | See Sweeney3 signature | | |

\* Signatures are named by first author and number of transcripts included in the model (e.g. Author11). Numbers in brackets indicate the original number of transcripts in the published model. Some signatures have a reduced number of transcripts when translated to RT-qPCR due to duplicate transcript symbols (IDs), high-primer probe failure rate (poor amplification efficiency), or where transcript sequences from an original discovery cohorts could not be mapped to a more recent reference transcriptome.
† The CERKL1 primer-probe assay failed during panel optimisation and was removed from the Duffy10 signature.
‡ Three transcripts (C1QB, C1QC, and GBP6) were excluded from the Kaforou27 signature due to a high primer-probe assay failure rate. Unique primer-probe assays could not be designed for both of the FCGR1B transcript variants; only one FCGR1B primer-probe was included in the Kaforou22 model. We were unable to design a primer-probe for LOC728744; all information had been withdrawn from both the NCBI and RefSeq databases.
LTBI, latent tuberculosis infection. OD, other diseases. HC, healthy controls. SVM, support vector machines. LASSO, Least Absolute Shrinkage and Selection Operator.

### Statistical analysis

All analyses were performed using R Statistical Software (v4.2.1). Signature score distributions were summarised using the median and interquartile range (IQR). Differences between two groups were assessed using the Wilcoxon Signed-Rank test for paired data and the Mann–Whitney U test for unpaired data. For comparisons across three or more groups, the Kruskal-Wallis test was used for unpaired data and the Friedman test for paired data. Spearman’s rank correlation was used to assess relationships between signature scores. Only samples with complete assay data were included in the correlation matrices. A two-sided alpha of <0.05 was considered statistically significant.

To evaluate prognostic performance, area under the receiver operating characteristic (ROC) curve (AUC) was calculated using the *pROC* package,^33^ and 95% confidence intervals (CIs) were computed using the DeLong method.^34^ AUCs were calculated for each candidate signature to distinguish cure from treatment failure, recurrence, or death at each treatment timepoint. Sensitivity, specificity, positive predictive value (PPV), and negative predictive value (NPV) were calculated using binary outcome indicators and standard formulae, and benchmarked against the WHO Target Product Profile (TPP) for TB treatment monitoring and optimisation (**Table S3**), at baseline/diagnosis (sensitivity 90%, specificity 70%), during (sensitivity 75%, specificity 80%), and at the end (sensitivity 80%, specificity 90%) of TB treatment.^35,36^ The number needed to treat (NNT) was calculated as 1/PPV. Wilson score intervals were used to calculate 95% CIs for performance metrics.

### Ethical approval

The study protocol was approved by the ethics committee of the Maternidade Climério de Oliveira in Salvador, Brazil; the institutional research ethics committees at each participating site; and Vanderbilt University Medical Center. The University of Cape Town Faculty of Health Sciences Human Research Ethics Committee (HREC 594/2018) approved the analysis of transcriptomic signatures on all samples collected. All participants provided written informed consent.

## RESULTS

### Recruitment of TB patients and matching

Between June 2015 and June 2019, 1,188 TB patients were enrolled in the RePORT-Brazil cohort (**Figure 1**), of whom 1,041 with culture-confirmed drug-susceptible pulmonary TB had baseline RNA samples available. Of the 1,041 participants eligible for this sub-study, 637 (61.2%) completed follow-up with recurrence-free cure; 297 (28.5%) did not complete treatment or were lost to follow-up; 28 (2.7%) died from TB-unrelated causes; and 13 (1.2%) discontinued follow-up for other reasons. Overall, 66 (6.3%) participants with an unfavourable treatment outcome were included in the analysis: 33 (3.2%) who failed treatment, 24 (2.3%) who died from TB-related or unknown causes during treatment or study follow-up, and 9 (0.9%) who had TB recurrence. Of the nine recurrence cases, three were microbiologically confirmed and six were clinically diagnosed.

**Figure 1.**
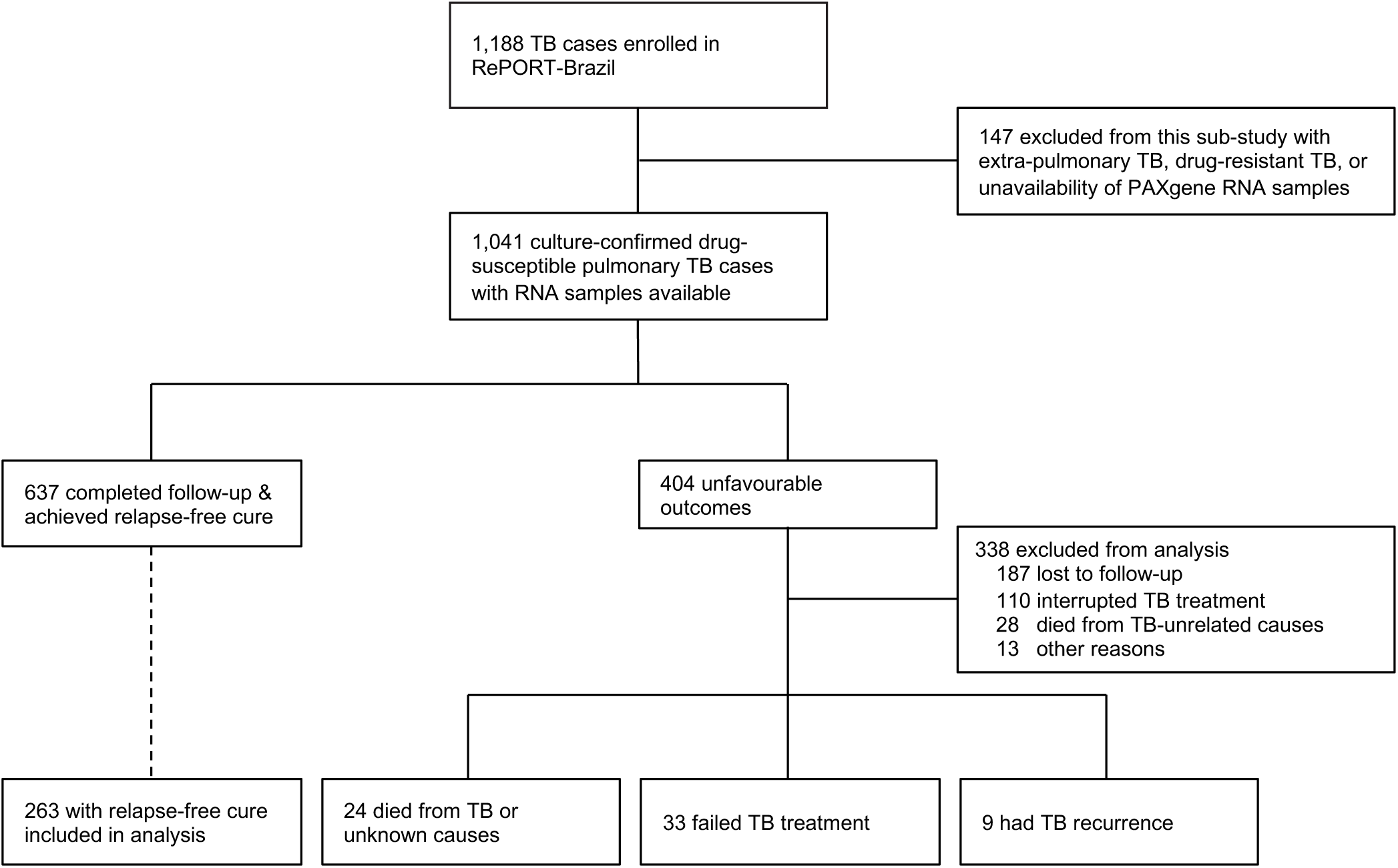
Study flow diagram.

All participants with an unfavourable outcome (treatment failure, death, or recurrence) were matched to 263 participants with recurrence-free cure (**Figure 1**). This included 173 participants selected in both matching procedures, 42 in the individual-endpoint matching only, and 48 in the combined-endpoint matching only. Their characteristics are displayed in **Table 2**, stratified by outcomes status.

**Table 2.**
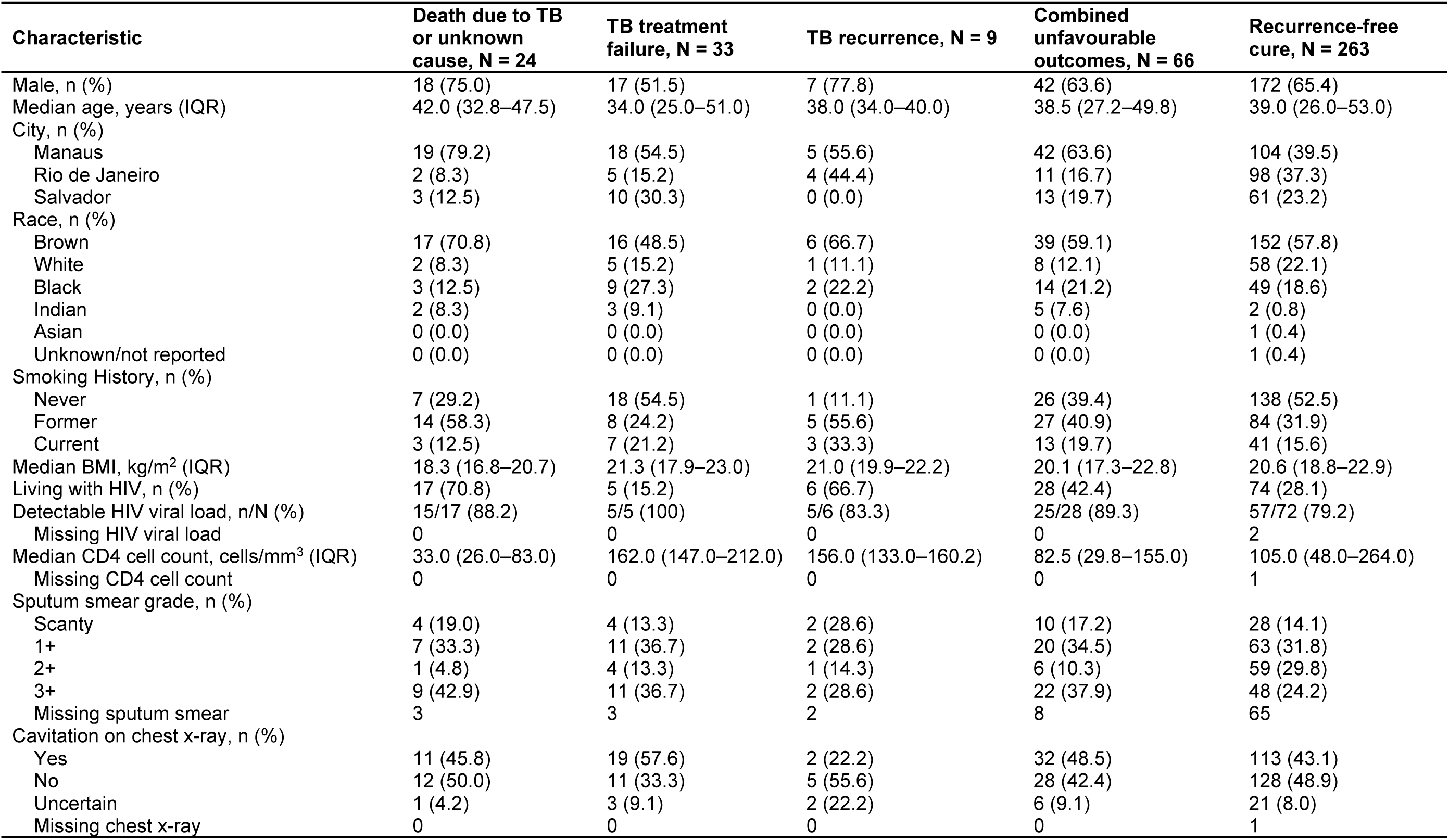
Characteristics of the study population.

| Characteristic | Death due to TB or unknown cause, N = 24 | TB treatment failure, N = 33 | TB recurrence, N = 9 | Combined unfavourable outcomes, N = 66 | Recurrence-free cure, N = 263 |
| --- | --- | --- | --- | --- | --- |
| Male, n (%) | 18 (75.0) | 17 (51.5) | 7 (77.8) | 42 (63.6) | 172 (65.4) |
| Median age, years (IQR) | 42.0 (32.8–47.5) | 34.0 (25.0–51.0) | 38.0 (34.0–40.0) | 38.5 (27.2–49.8) | 39.0 (26.0–53.0) |
| City, n (%) |  |  |  |  |  |
| Manaus | 19 (79.2) | 18 (54.5) | 5 (55.6) | 42 (63.6) | 104 (39.5) |
| Rio de Janeiro | 2 (8.3) | 5 (15.2) | 4 (44.4) | 11 (16.7) | 98 (37.3) |
| Salvador | 3 (12.5) | 10 (30.3) | 0 (0.0) | 13 (19.7) | 61 (23.2) |
| Race, n (%) |  |  |  |  |  |
| Brown | 17 (70.8) | 16 (48.5) | 6 (66.7) | 39 (59.1) | 152 (57.8) |
| White | 2 (8.3) | 5 (15.2) | 1 (11.1) | 8 (12.1) | 58 (22.1) |
| Black | 3 (12.5) | 9 (27.3) | 2 (22.2) | 14 (21.2) | 49 (18.6) |
| Indian | 2 (8.3) | 3 (9.1) | 0 (0.0) | 5 (7.6) | 2 (0.8) |
| Asian | 0 (0.0) | 0 (0.0) | 0 (0.0) | 0 (0.0) | 1 (0.4) |
| Unknown/not reported | 0 (0.0) | 0 (0.0) | 0 (0.0) | 0 (0.0) | 1 (0.4) |
| Smoking History, n (%) |  |  |  |  |  |
| Never | 7 (29.2) | 18 (54.5) | 1 (11.1) | 26 (39.4) | 138 (52.5) |
| Former | 14 (58.3) | 8 (24.2) | 5 (55.6) | 27 (40.9) | 84 (31.9) |
| Current | 3 (12.5) | 7 (21.2) | 3 (33.3) | 13 (19.7) | 41 (15.6) |
| Median BMI, kg/m<sup>2</sup> (IQR) | 18.3 (16.8–20.7) | 21.3 (17.9–23.0) | 21.0 (19.9–22.2) | 20.1 (17.3–22.8) | 20.6 (18.8–22.9) |
| Living with HIV, n (%) | 17 (70.8) | 5 (15.2) | 6 (66.7) | 28 (42.4) | 74 (28.1) |
| Detectable HIV viral load, n/N (%) | 15/17 (88.2) | 5/5 (100) | 5/6 (83.3) | 25/28 (89.3) | 57/72 (79.2) |
| Missing HIV viral load | 0 | 0 | 0 | 0 | 2 |
| Median CD4 cell count, cells/mm<sup>3</sup> (IQR) | 33.0 (26.0–83.0) | 162.0 (147.0–212.0) | 156.0 (133.0–160.2) | 82.5 (29.8–155.0) | 105.0 (48.0–264.0) |
| Missing CD4 cell count | 0 | 0 | 0 | 0 | 1 |
| Sputum smear grade, n (%) |  |  |  |  |  |
| Scanty | 4 (19.0) | 4 (13.3) | 2 (28.6) | 10 (17.2) | 28 (14.1) |
| 1+ | 7 (33.3) | 11 (36.7) | 2 (28.6) | 20 (34.5) | 63 (31.8) |
| 2+ | 1 (4.8) | 4 (13.3) | 1 (14.3) | 6 (10.3) | 59 (29.8) |
| 3+ | 9 (42.9) | 11 (36.7) | 2 (28.6) | 22 (37.9) | 48 (24.2) |
| Missing sputum smear | 3 | 3 | 2 | 8 | 65 |
| Cavitation on chest x-ray, n (%) |  |  |  |  |  |
| Yes | 11 (45.8) | 19 (57.6) | 2 (22.2) | 32 (48.5) | 113 (43.1) |
| No | 12 (50.0) | 11 (33.3) | 5 (55.6) | 28 (42.4) | 128 (48.9) |
| Uncertain | 1 (4.2) | 3 (9.1) | 2 (22.2) | 6 (9.1) | 21 (8.0) |
| Missing chest x-ray | 0 | 0 | 0 | 0 | 1 |

### Transcriptomic signatures track response to TB treatment

We investigated whether host blood transcriptomic signatures could track treatment response in pulmonary TB by measuring scores at multiple timepoints during treatment. Baseline scores were available for 328 of 329 participants; one sample from a participant with treatment failure was missing. As shown in **Figure S1**, most signatures were moderately to strongly correlated with each other (Spearman *ρ* ranging from 0.39 to 0.96), suggesting a shared underlying biological signal. Given the strong inter-correlation, we selected Thompson5 and XpertHR as representative examples for figures and analyses because (1) their performance as response biomarkers varied by timepoint, and (2) the Thompson5 signature was the first published parsimonious treatment response signature, while XpertHR has been adapted for a cartridge-based point-of-care assay. Notably, Duffy4—a signature of protection from TB in a contained Mtb mouse model^37^—was less correlated with other signatures (*ρ* 0.14–0.57), likely due to its predominant mapping to T-cell gene pathways.

We next evaluated signature trajectories over time. Thompson5 and XpertHR (**Figure 2A**) were highest at baseline and declined progressively during treatment, despite substantial inter-individual variability. These downward trends were confirmed by Friedman and Jonckheere–Terpstra tests (p<1×10⁻^54^) and consistent negative correlations over time (Kendall’s τ=−0.47 for both). Both signatures also showed strong discrimination between treatment stages. They readily distinguished baseline from month 2 samples (AUC≥0.75) and from EoT samples (AUC=0.91; **Figure 2B**). To explore whether these patterns extended across the broader set of transcriptomic signatures, we compared AUCs for all 22 signatures across timepoints **(Figure 2C).** Most signatures showed moderate discrimination between baseline and month 2, weaker separation between month 2 and EoT, and strong discrimination for baseline and EoT (**Figure 2C**).

**Figure 2.**
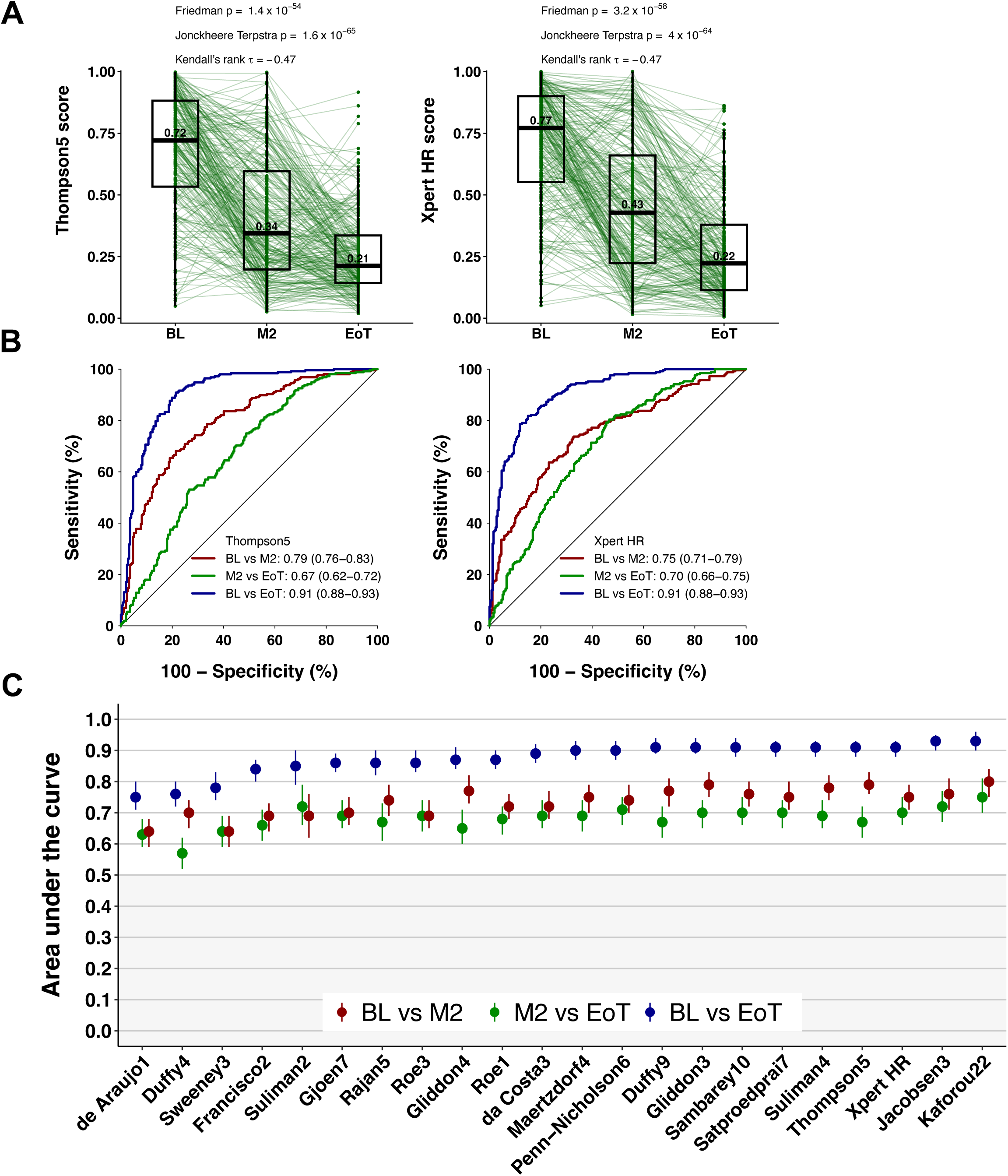
Transcriptomic signatures track response to TB treatment. (A) Line graphs depicting score trajectories of two representative signatures, Thompson5 (left) and Xpert Host-Response (HR; right), measured at start (BL=baseline), month 2 (M2), and end of treatment (EoT) for drug-susceptible TB among participants with recurrence-free cure. Each line represents a participant. Boxes depict the IQR, the midline represents the median (shown), and the whiskers indicate the IQR ± (1.5 × IQR). (B) Thompson5 (left) and XpertHR (right) signature receiver operating characteristic (ROC) curves with area under the curve (AUC) estimates and 95% CIs for differentiating paired signature scores measured at the BL versus M2 of treatment (red), M2 versus EoT (green), and BL versus EoT (blue). (C) Summary of ROC performance (AUC) for differentiating paired signature scores measured at the BL versus M2 of treatment (red), M2 versus EoT (green), and BL versus EoT (blue). Signatures are ordered by AUC estimates for BL versus EoT. Circles indicate the AUC estimate and error bars indicate 95% CIs.

### Signatures predicted unfavourable treatment outcomes

Next, we assessed the performance of transcriptomic signatures to differentiate between individuals who achieved clinical cure and remained disease free during follow-up, and those who experienced unfavourable treatment outcomes. Median signature scores for Thompson5 and XpertHR were lower in all timepoints among individuals with cure, relative to individuals with poor treatment outcomes (**Figure 3A-B**). Scores for both signatures were significantly higher in individuals who ultimately developed recurrent TB compared to those who achieved cure at baseline, and at EoT for XpertHR, but not Thompson5. Consistent with this, XpertHR and Thompson5 predicted recurrence at baseline and month 2, while XpertHR also predicted recurrence at EoT (**Figure 3C; Table S4**). Interestingly, relative to EoT, signature scores increased further at the time of recurrent TB diagnosis, especially for the Thompson5 signature (**Figure 3A-B**). Similar significant prediction of recurrent TB was obtained for virtually all signatures tested, suggesting strongly elevated expression of IFN-response and inflammatory genes among participants at risk of recurrence (**Figure 3D; Table S4**).

**Figure 3.**
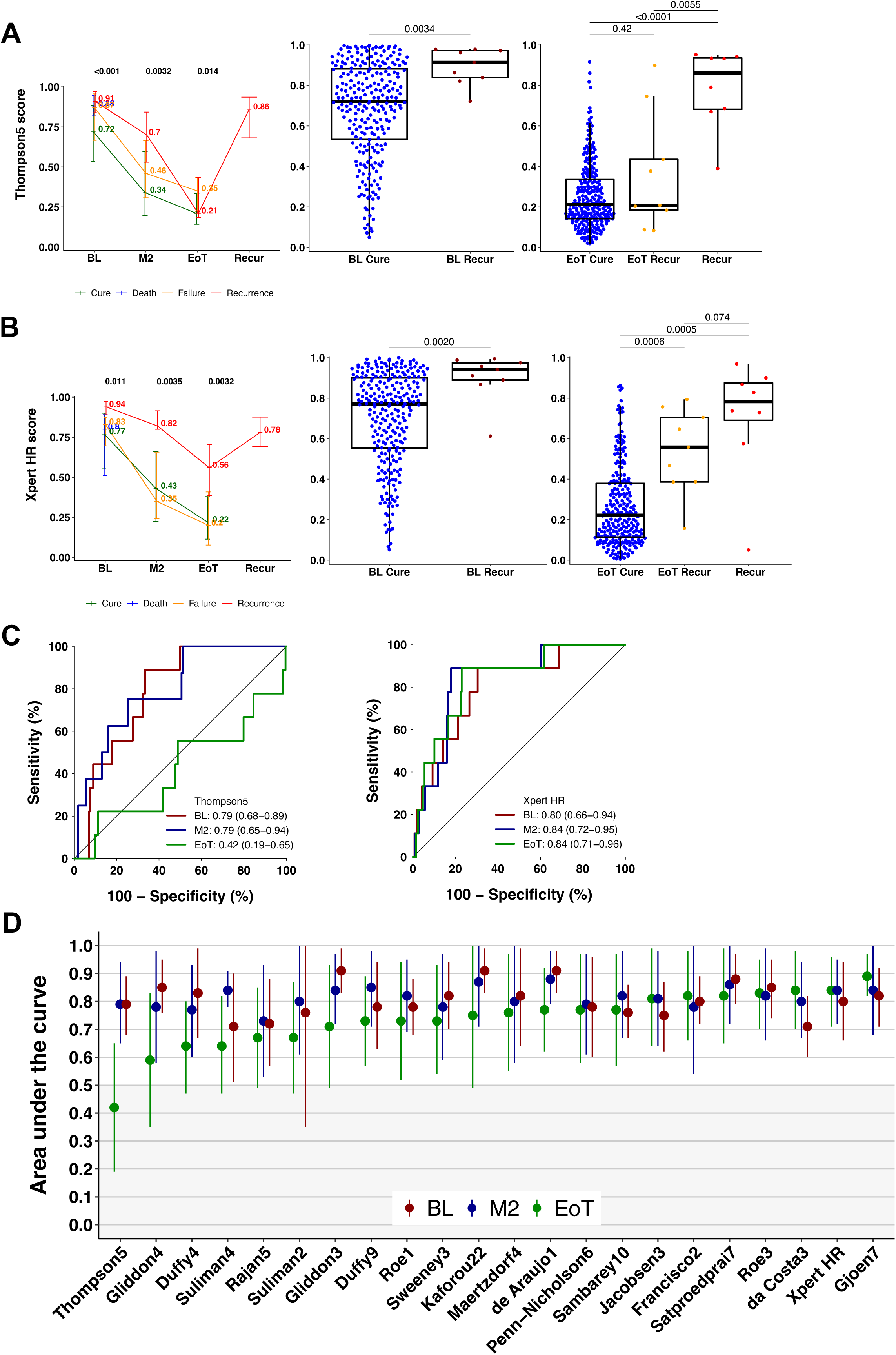
Transcriptomic signatures distinguish individuals with TB recurrence from those with recurrence-free cure. (A–B) Longitudinal changes for two representative signatures, Thompson5 (A) and XpertHR (B), among participants with recurrence-free cure (green), recurrence (red), failure (orange), and death (blue). Left panels show median scores (error bars indicate interquartile range) at baseline (BL), month 2 (M2), end of treatment (EoT), and at time of recurrent TB diagnosis (Recur). Middle panels compare scores at BL between those with cure (BL Cure) vs. recurrence (BL Recur). Right panels show EoT scores between those with cure (EoT Cure) vs. recurrence (EoT Recur), and scores at the time of recurrence (Recur). P values of between-group differences were computed by Wilcoxon tests. (C) Receiver operating characteristic (ROC) curves evaluating Thompson5 (left) and XpertHR (right) for prediction of recurrence at BL (red), M2 (blue), and EoT (green). Area under the curve (AUC) estimates and 95% CIs are shown. (D) Summary of ROC performance (AUC) for all transcriptomic signatures at BL (red), M2 (blue), and EoT (green), for predicting recurrence. Signatures are ordered by AUC estimates at EoT. Circles indicate the AUC estimate and error bars indicate 95% CIs.

When benchmarked against the WHO TPP for TB treatment monitoring and optimisation (**Table S3**), 2 of 22 transcriptomic signatures met the minimum prognostic criteria for predicting recurrence at baseline (90% sensitivity, 70% specificity), 13 signatures met the benchmark at month 2 (75% sensitivity, 80% specificity), and none achieved the benchmark at EoT (80% sensitivity, 90% specificity; **Table S5**). An additional 14 of the 22 signatures had 95% confidence interval upper limits that exceeded the benchmark at baseline, 12 signatures at month 2, and 7 at EoT. The best-performing signature at the EoT for prediction of recurrence was Gjøen7, which achieved a sensitivity of 66.7% (95% CI 35.4–87.9), meaning that approximately two-thirds of recurrences would be detected, with specificity fixed at 90%. Sensitivity fell short of the 80% WHO TPP minimum target. The corresponding PPV was 18.2% (95% CI 8.6–34.4) and NPV 98.7% (95% CI 96.3–99.6), at a recurrence rate of 3.4%. The NNT was 5.5 (95% CI 2.9–11.6), indicating that treatment would need to be prolonged in about six patients with elevated scores to potentially prevent one recurrence. At month 2, XpertHR had a sensitivity of 88.9% (95% CI 56.5–98.0) with specificity fixed at 80%, exceeding the WHO TPP, with PPV 13.1% (95% CI 6.8–23.8), NPV 99.5% (95% CI 97.3–99.9), and NNT 7.6 (95% CI 4.2–14.7). At baseline, Gjøen7 had a sensitivity of 88.9% (95% CI 56.5–98.0), approaching the WHO TPP with specificity fixed at 70%, PPV 9.3% (95% CI 4.8–17.3), NPV 99.0% (95% CI 96.6–99.7), and NNT 10.8 (95% CI 5.8–20.9).

We also investigated the performance of transcriptomic signatures for treatment failure. However, the discriminatory performance to predict failure was weak across all timepoints. Thompson5 significantly differentiated between those with treatment failure and cure, but with AUCs not exceeding 0.70 (**Figure 4A**). The XpertHR signature did not differentiate between those with treatment failure and cure at any timepoint (**Figure 4B**), a result that was observed for most signatures assessed (**Figure 4C; Table S6**). All signatures fell well short of the WHO TPP prognostic criteria at all treatment timepoints (**Table S7**).

**Figure 4.**
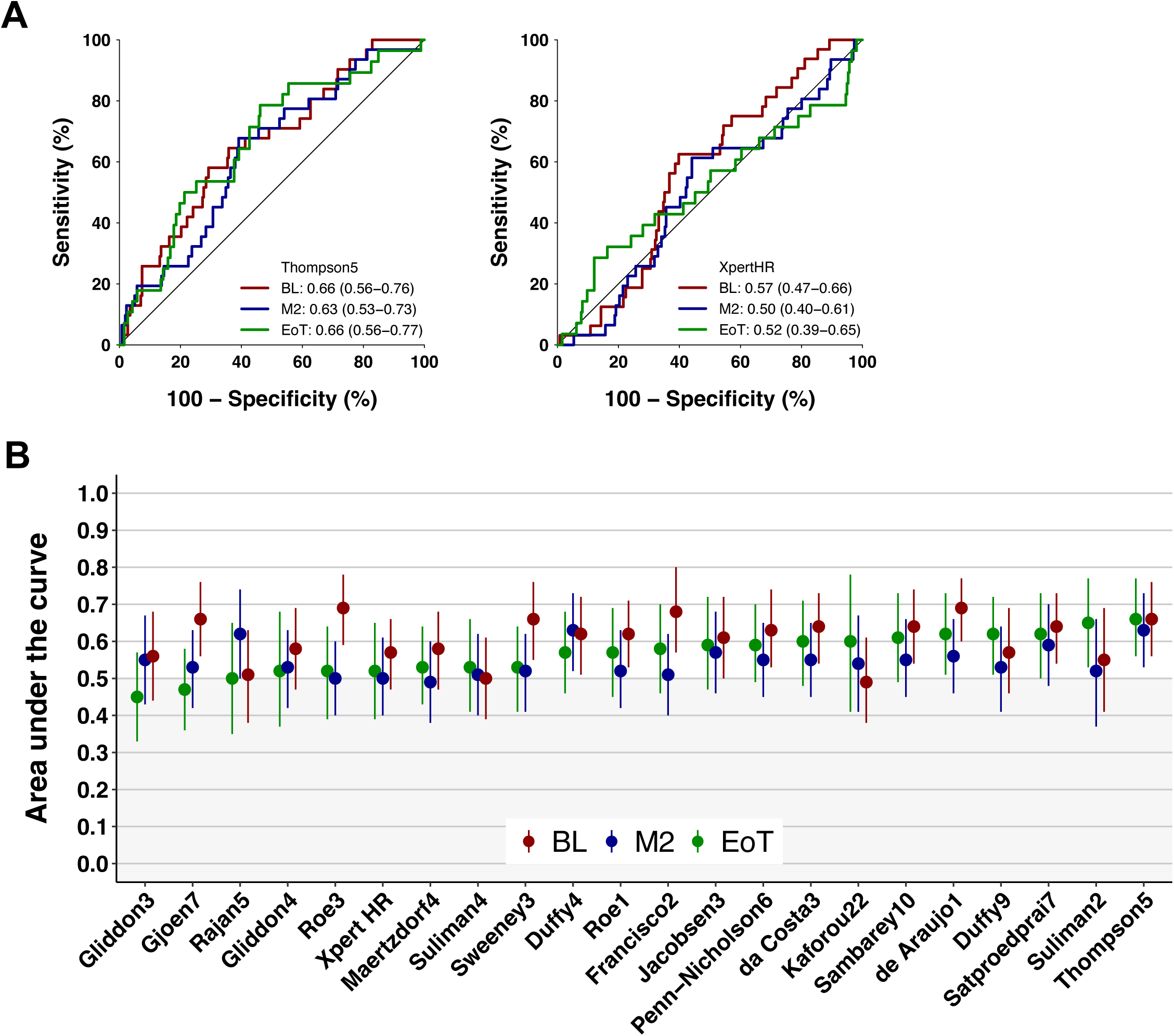
Transcriptomic signature performance for prediction of treatment failure. (A) Receiver operating characteristic (ROC) curves for two representative signatures (left: Thompson5; right: XpertHR), comparing individuals who experienced treatment failure versus those with recurrence-free cure, at baseline (BL, red), month 2 (M2, blue), and end of treatment (EoT, green). Area under the curve (AUC) estimates and 95% CIs are shown. (B) Summary ROC performance (AUC) for all transcriptomic signatures at BL (red), M2 (blue), and EoT (green), for predicting treatment failure. Signatures are ordered by AUC estimates at EoT. Circles indicate the AUC estimate and error bars indicate 95% CIs.

Finally, we determined if transcriptomic signatures could predict death due to TB or unknown cause in this cohort. At baseline, the Thompson5 signature distinguished individuals who later died from those who achieved recurrence-free cure (**Figure 5A, left**). In contrast, the XpertHR signature demonstrated no discrimination (**Figure 5A, right**). When assessed across the full set of signatures at baseline, 11 other signatures also showed significant differentiation between those who ultimately died and controls who remained disease-free (**Figure 5B; Table S8**). No signature met the WHO TPP at baseline (90% sensitivity, 70% specificity) for prediction of death (**Table S9**), but Thompson5 came closest with sensitivity of 84.2% (95% CI 62.4–94.5) at fixed 70% specificity.

**Figure 5.**
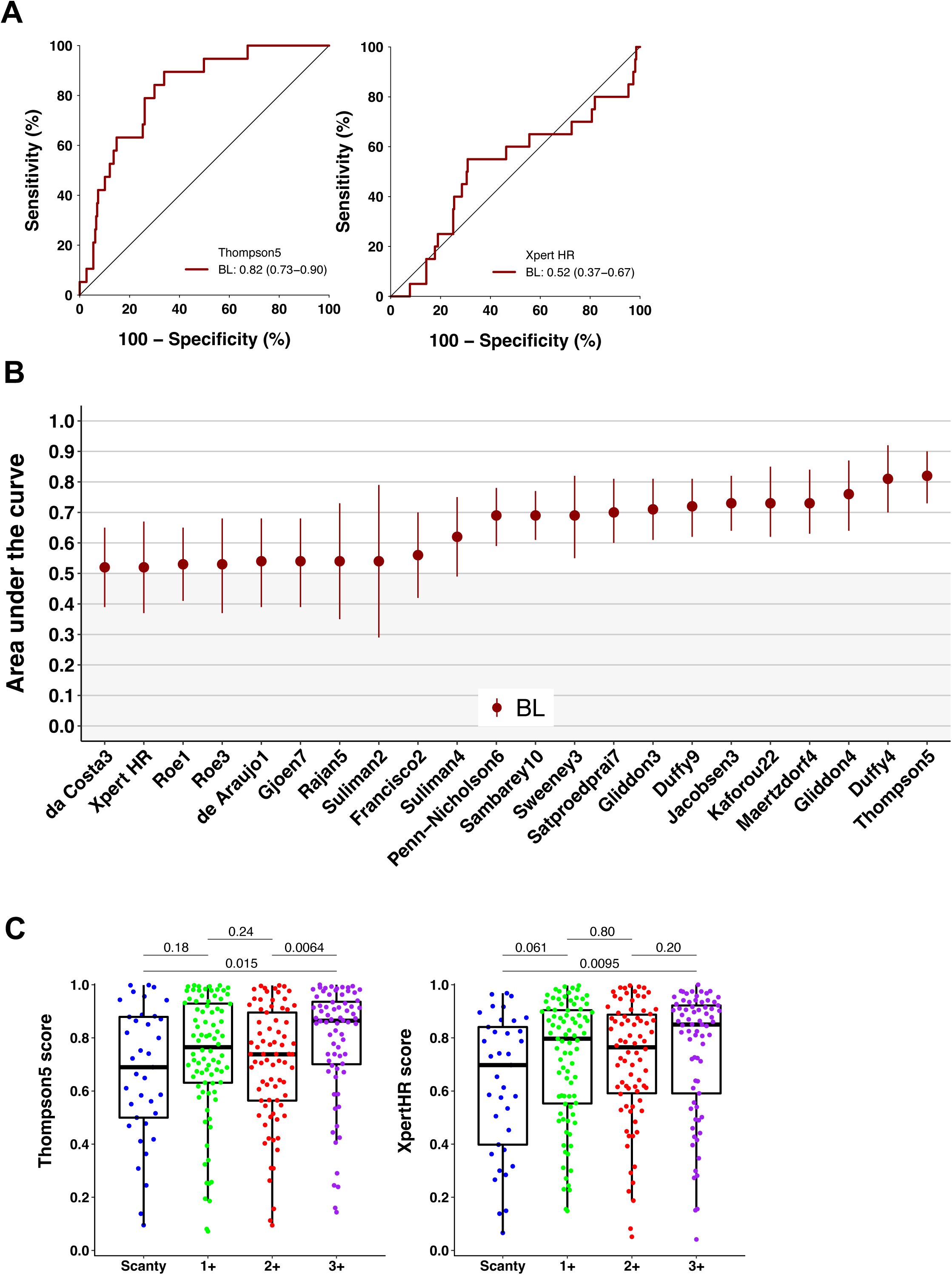
Transcriptomic signatures predict TB-related or unknown-cause death and are associated with sputum bacillary burden. (A) Receiver operating characteristic (ROC) curves for two representative signatures (left: Thompson5; right: XpertHR) at baseline (BL) comparing individuals who died during or after treatment to those who achieved recurrence-free cure. Area under the curve (AUC) estimates and 95% CIs are shown. (B) Summary of receiver operating characteristic (ROC) performance (AUC) for all transcriptomic signatures at BL (red) for prediction of TB-related or unknown-cause death. Signatures are ordered by AUC estimates at BL. Circles indicate the AUC estimate and error bars indicate 95% CIs. (C) Boxplots of baseline Thompson5 (left) and XpertHR (right) scores stratified by sputum smear microscopy grade (scanty, 1+, 2+, 3+). P values correspond to pairwise comparisons (Wilcoxon rank-sum tests). Each dot represents a participant. Boxes depict the IQR, the midline represents the median, and the whiskers indicate the IQR ± (1.5 × IQR).

To explore whether elevated transcriptomic scores reflected higher bacterial burden, we stratified Thompson5 and XpertHR scores by sputum smear grade at baseline. Although scores for many signatures, including Thompson5 and XpertHR, were significantly higher in those with smear microscopy scores of 3+ compared to scanty (**Figure 5C**), the proportions of participants with pulmonary cavities or other commonly used measures of disease severity did not differ. We then performed a multivariable linear regression analysis to identify factors associated with baseline signature scores (**Table S10**). Independent of treatment outcome, a higher sputum smear grade was associated with elevated baseline scores in 13/22 signatures. No other clinical variables, including HIV status and cavitation, showed consistent associations across signatures.

## DISCUSSION

We evaluated previously published, parsimonious host-blood transcriptomic signatures for their ability to track treatment response and predict recurrent TB, treatment failure, or death among adults with pulmonary TB in Brazil. No validated non-sputum biomarkers currently exist to monitor response or guide regimen choice and duration. To our knowledge, this is the first comprehensive RT-qPCR evaluation of these signatures in a large, well-characterised prospective cohort, reflecting the heterogeneity of real-world treatment response.

Multiple host-blood transcriptomic signatures exhibited dynamic, treatment-responsive trajectories, with scores generally declining over therapy, consistent with resolution of systemic inflammation and immune dysregulation in clinical TB.^18,38,39^ Despite substantial inter-individual heterogeneity, the signatures discriminated between samples collected at baseline, month 2, and the EoT. Our findings are consistent with previous studies showing treatment-associated shifts in whole-blood gene expression.^11,12,15,16,38,40^ However, whether such signatures can prospectively identify patients at high risk of poor outcomes has remained uncertain.

In our cohort, several parsimonious signatures, most originally developed for TB diagnosis or prognosis, predicted TB recurrence. Additionally, several met the WHO TPP minimum criteria at baseline or month 2, supporting their potential for individualised, biomarker-guided treatment optimisation. Such tools could prompt longer treatment for high-risk patients. Conversely, such tools may justify shortening in early responders whose scores normalise towards healthy ranges (consistent with evidence that cure is achievable with shorter regimens^7,8^), thereby reducing drug exposure and improving adherence and optimising resource use.

Several signatures also predicted death, indicating that blood-based biomarkers could help identify patients at heightened risk who may benefit from more intensive assessment and care. In our cohort, most deaths occurred among PLHIV (70.8%; median CD4 33 cells/mm^3^, IQR 26–83), disproportionate to their representation among participants with recurrence-free cure (28.1%; median CD4 105 cells/mm^3^, IQR 48–264; **Table 2**), which may partly explain the strong association between inflammatory activity and mortality. This aligns with findings in HIV-associated TB, where elevated inflammatory proteins predict death.^41^

Notably, the signatures poorly predicted treatment failure across timepoints; this may reflect (i) the predominantly focal nature of bacteriological failure, due to bacilli persisting in poorly perfused lesions with limited systemic inflammatory signal; (ii) etiologic heterogeneity (suboptimal adherence, pharmacokinetic variability, drug–drug interactions, or undetected resistance) that is not encoded in host transcriptomic activity; and (iii) the relatively small number of failures, reducing statistical power. Collectively, these observations suggest that blood signatures index global inflammatory disease activity—relevant to recurrence and death—better than they capture pharmacologic or microbiologic determinants of early bacteriological failure. However, we note that previous studies have reported transcriptomic signatures that could differentiate between individuals with treatment cure and those with treatment failure.^12^

Transcriptomic scores correlated with bacillary burden, increasing with higher sputum smear grade. Although not uniform across all signatures, this pattern suggests that host-response signatures may partially reflect disease severity. Previous reports have demonstrated that the Sweeney3 and Penn-Nicholson6 signatures correlated with the extent of pulmonary disease quantified by PET-CT imaging.^13,14^ We observed no associations with cavitation, underscoring that these signals capture systemic inflammatory activity not fully represented by structural radiographic markers.

Our study had limitations. Recurrence events were few and two-thirds were not microbiologically confirmed, resulting in wide confidence intervals and reducing certainty of endpoint ascertainment, which potentially affected estimates of biomarker performance. The absence of culture confirmation and genotyping also prevented linkage of recurrence isolates to the index strain, precluding classification of recurrence as relapse or reinfection, outcomes with distinct biology that may differentially influence host-response signatures. Death due to TB or unknown cause and symptom-triggered (rather than scheduled) post-treatment recurrence assessments may have led to outcome misclassification and under-ascertainment of asymptomatic recurrence. Our analyses were restricted to a risk-score–matched subset, which excluded participants who were lost to follow-up. As a result, diagnostic estimates reflect the matched analytic sample and may not generalise to the full RePORT-Brazil cohort; PPV/NPV and NNT in particular are not prevalence-representative. Despite propensity matching, we did not match on all prognostically important characteristics (e.g., cavitation, smear grade), and residual confounding by measured and unmeasured factors may persist. To mitigate overfitting, we used a previously optimised RT-qPCR protocol, calculated signature scores with locked-down models without re-training on the present cohort, and remained blinded to clinical phenotypes throughout. Nevertheless, we evaluated 22 signatures and highlighted those performing best in this dataset; generalisability to other settings and populations remains to be established.

In summary, parsimonious host-blood transcriptomic signatures show promise as biomarkers to monitor treatment response and to prognosticate poor outcomes in pulmonary TB. A necessary next step is to validate that such signatures predict relapse or failure in the setting of treatment-shortening regimens (e.g., the 4-month rifapentine–moxifloxacin regimen from TBTC Study 31/ACTG A5349),^7^ using retrospective analyses or prospectively embedded studies. Contingent on positive validation, a pragmatic biomarker-guided trial, assigning longer or shorter regimens on the basis of pre-specified high/low signature thresholds would then be warranted to establish clinical utility and safety.^42^ An optimal test would be non-sputum (ideally peripheral/finger-prick blood), accurately reflect disease activity and bacillary clearance across therapy, and be especially valuable where sputum is hard to obtain. Used alongside standard clinical and microbiological assessment, such signatures could facilitate precision care by supporting regimen-shortening for early responders and targeted intensification for high-risk patients (e.g., additional drugs or extended treatment duration). Our results provide a rationale and candidate signatures for advancing these biomarkers into pragmatic trials of individualised TB treatment.

## AUTHORS’ CONTRIBUTIONS

SCM, BBA, TRS, and TJS conceived the idea. SCM, BBA, TRS, and TJS raised funds and/or provided the resources. BBA, ALK, MC-S, and VCR were responsible for all site-level activities, including recruitment, clinical management, and data collection. SCM, AMSA, MF, and MCF provided operational, technical, laboratory support, or project management. VMM contributed to transcriptomic signature panel design. ME processed samples and performed the experiments. SCM, GCA and MCF verified the underlying data. SCM analysed the data. SCM, MAP, TRS, and TJS interpreted the results and wrote the first draft. All authors had full access to the data, confirm the integrity of the data and its presentation, agree with its interpretation as discussed in the manuscript, and reviewed, revised, and approved the manuscript before submission. The corresponding author had final responsibility for the decision to submit for publication.

## DATA AVAILABILITY STATEMENT

Deidentified qPCR data, signature scores, and TB endpoint data underlying the results have been deposited in Zivahub doi.org/10.25375/uct.30121768), an open access data repository hosted by the University of Cape Town’s institutional data repository powered by Figshare for Institutions. Additional clinical metadata are available on request from the RePORT Consortium (reportinternational.org).

