## Supplement for "Blood transcriptomic signatures predict poor treatment outcomes in drug-susceptible pulmonary TB in Brazil"

*RePORT-South Africa Consortium*
Nicole Bilek
Yolundi Cloete
Katie Hadley
Rieyaat Hassiem
Lungisa Jaxa
Stanley Kimbung Mbandi
Faheemah Meyer
Humphrey Mulenga
Onke Nombida
Alison September
Ashley Veldsman

*RePORT-Brazil Consortium*
Brenda Carvalho
Adriano Gomes
Cody Staats
Megan Turner

#### Supplemental Methods

##### Propensity (or risk) scores

Propensity (or risk) scores[1] were estimated using logistic regression. Age was included as a continuous covariate in the propensity/risk score model, while sex, HIV status, and site of enrolment were used as exact matching variables. Matching was performed using nearest-neighbour matching on the estimated propensity/risk score, with a caliper of 0.25—that is, participants with unfavourable outcomes were only matched to those with favourable outcomes whose propensity scores were within 0.25 standard deviations. Two matching procedures were conducted. First, participants with recurrence-free cure were matched, without replacement, at a 3:1 ratio to each subgroup with unfavourable outcomes (treatment failure, recurrence, and death) to enable outcome-specific contrasts. Second, a 3:1 matching was performed between participants with favourable outcomes and those with any combined unfavourable outcome to increase power and yield a single overall estimate comparable to composite endpoints used in TB studies. All matched participants from both procedures were included in the analysis.

##### Transcriptomic signature panel design and measurement

RNA plates were thawed in batches, cDNA synthesised with EpiScript reverse transcriptase (Lucigen, USA), and genes of interest pre-amplified using pools of TaqMan primer-probe assays (Thermo Fisher Scientific, USA; **Table S1**). Gene expression (raw cycle threshold, Ct) was quantified by microfluidic multiplex real-time quantitative PCR (RT-qPCR) using Standard BioTools (formerly Fluidigm, USA) 96.96 (96 samples multiplexed with 96 primer-probe assays) Gene Expression chips on a BioMark HD instrument (Standard BioTools). Quality control filters, batch correction, and calculation of the signature scores were performed using a pre-defined R script.

##### Data quality control

Gene expression data were analysed using a pre-defined R script with quality control filters that assessed the integrity and reproducibility of each chip. The following parameters were applied for extracting Ct values: Linear (Derivative) baseline correction, Quality Threshold of 0.3, and Auto (Global) for Ct Threshold Method using Biomark software version 4.5.2. No-template (water) and internal positive control samples were run on each chip. Chips with marked deviation (>5 standard deviations, Spearman correlation < 0.98, or concordance correlation coefficient < 0.95) in the internal positive control sample primer-probe assay raw Ct values or Penn-Nicholson6 (RISK6) signature score versus 11 historical runs, with amplification (raw Ct < 35 cycles) for any assay in the no-template control, or with more than 10% failed primer-probes, were repeated. Individual samples with more than 20% failed primer-probe reactions were classified as failed and no signature scores were computed. If less than 20% of primer-probe reactions failed for an individual sample, signature scores were computed where possible and signatures with missing primer-probe raw Ct values were deemed failed for that sample. Samples and primer-probe assays were run in singlet, and failed signature results for individual samples were assumed to follow a random distribution, thus not repeated, and excluded from analysis.

**Signature score calculation**

Signature scores were calculated from raw Ct measurements using the original algorithms developed for each individual signature, where possible (**Table 1**). Signatures discovered using methods other than RT-qPCR, such as RNA-sequencing or microarray, were subsequently re-parameterised to RT-qPCR.[2,3] In cases where predesigned TaqMan primer-probe assays were not available from the original publication, we used either inventoried primer-probes or designed novel TaqMan assays based on the published Illumina probe, RNA-seq transcript, or primer-probe sequence data. Furthermore, we contacted the authors of relevant publications when the required information was not available. We included three reference primer-probes (ACTR3, TMBIM6, and USF2; **Table S1**) for standardisation of gene expression for signatures which require a normalised (delta) Ct.

**Signature reparameterisation**

Eight signatures (da Costa3, Duffy9, Francisco2, Gjøen7, Jacobsen3, Roe3, Sambarey10, and Satproedprai7) developed using gene expression analysis techniques other than RT-qPCR required reparameterisation in order to compute signature scores from microfluidic multiplex RT-qPCR BioMark HD readouts. This was necessary because different platforms each have unique dynamic ranges and quantitative scales for quantifying gene expression levels. The machine learning models, and statistical methods used for computing signature scores therefore needed to be reconstructed (reparameterised) using Ct values generated from RT-qPCR data on the BioMark HD platform. This ensured that there was uniformity in the way scores were calculated for different signatures and how results were interpreted. The model parameters for the eight signatures were reconstructed using the original published methods[4-11] to best differentiate between TB cases and controls when adapted to Ct values, generated using PAXgene RNA samples from a previously described cross-sectional TB cohort (CTBC)[12,13]. This cohort is completely independent of the RePORT-Brazil study and included both HIV positive and HIV negative TB cases and healthy controls; use of this independent dataset for reparameterisation reduced potential bias. The RT-qPCR protocol for the reparameterisation experiments was consistent with the methods used for all RT-qPCR in this paper.

Model training was done on individuals who were HIV negative (59 LTBI; 50 TB). 70% of HIV negative individuals (42 LTBI; 34 TB) made up the training set and the remaining 30% comprised the test set (17 LTBI; 16 TB). Once a model constructed on the training set demonstrated discrimination on the test set, then both sets were combined to reparameterise on the entire dataset (**Table S2**). Receiver operating characteristic (ROC) curves were generated to validate the performance of each model's ability to separate HIV-positive TB cases (n=39) from HIV-positive LTBI controls (n=36) (**Table S2**).

A number of signatures had not previously been evaluated using the Biomark RT-qPCR instrument, but did not require reparameterisation even though they were discovered using approaches other than RT-qPCR. Examples include the Gliddon3/4,[14] Kaforou25,[15] and Rajan5[16] signatures. Gliddon3/4 and Kaforou25 were discovered using microarray data using a simple Disease Risk Score (DRS), which sums normalized expression values from all upregulated transcripts and subtracts the normalized expression values of all downregulated transcripts, making the signature essentially “self-normalizing”. Rajan5 was also discovered based on microarray gene expression data, but developed using the signed and unsigned sums methods, which also allowed the signature to be “self-normalising”.

##### Batch correction and score normalisation

Batch correction was performed to correct for differences in signature score distribution between microfluidic chips using a quantile regression method with the adjust_batch function in the batchtma R package.[17] This method unifies lower quantiles (25th percentile) and ranges between the lower and upper quantile (75th percentile) between batches, allowing for differences in both parameters due to confounders. Thereafter, signature scores were winsorized (clipped) to account for extreme outliers (Winsorize function, DescTools R package[18]), log transformed to normalise score distribution (log function, base R package), converted to z-scores (scale function, base R package), and compared using the cumulative distribution function (pnorm function, stats R package). De Araujo1, Rajan5, and Roe1 signatures were multiplied by −1 to obtain a positive correlation for all signatures.

##### Blinding

Participants and study staff responsible for TB investigation in Brazil were blinded to transcriptomic signature scores. Transcriptomic signature scores were measured by laboratory personnel in South Africa who were blinded to participant treatment outcome. Signature scores and TB microbiology results were maintained in different files, which were integrated only after the study database had been cleaned and locked and the RT-qPCR data was cleaned and finalised.

#### Supplemental Figures

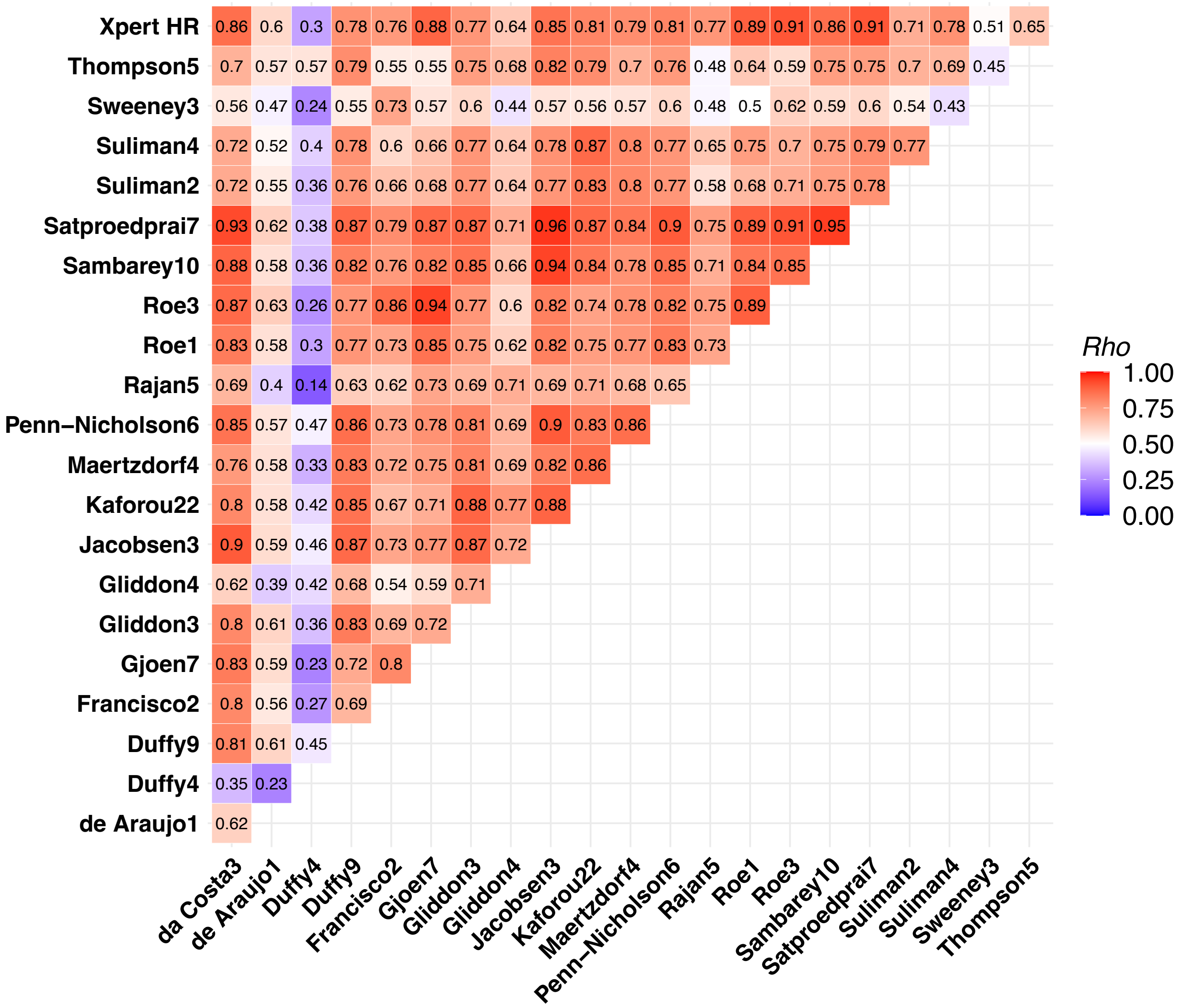

**Figure S1.** Correlation between signature scores.

Signature score correlation matrix with the Spearman-rank-order correlation coefficients (*rho*) among samples from TB patients, at all timepoints (n= 403 #). De Araujo1, Rajan5, and Roe1 signatures were multiplied by −1 to obtain a positive correlation for all signatures.

#Only participants with scores available for all signatures were included in the Spearman correlation analysis. Xpert HR=Xpert Host-Response.

#### Supplemental Tables

**Table S1.** TaqMan PCR primer-probe panel for the 96.96 gene expression integrated fluidic circuit.

| Gene Symbol | TaqMan Assay ID | Assay excluded** | Reference probe | # of signatures | da Costa3 [4] | de Araujo1 [19] | Duffy4 (5)# [20] | Duffy9 (10)† [5] | Francisco2 [6] | Gjøen7 [7] | Gliddon3 [14] | Gliddon4 [14] | Jacobsen3 [8] | Kaforou22 (27)‡ [15] | Maertzdorf4 [21] | Penn-Nicholson6 [13] | Rajan5 [16] | Roe1 [22] | Roe3 [9] | Sambarey10 [10] | Satproedprai7 [11] | Suliman2 [23] | Suliman4 [23] | Sweeney3 [24,25] | Thompson5 [26] | Xpert HR [27] |
|---|---|---|---|---|---|---|---|---|---|---|---|---|---|---|---|---|---|---|---|---|---|---|---|---|---|---|
| ACTR3 | Hs01029159_g1 | | X | 15 | X | X | X | X | X | X | X | X | X | X | | | X | X | X | X | X | | | | | X |
| TMBIM6 | Hs00162661_m1 | | X | 15 | X | X | X | X | X | X | X | X | X | X | | | X | X | X | X | X | | | | | X |
| USF2 | Hs01100994_g1 | | X | 15 | X | X | X | X | X | X | X | X | X | X | | | X | X | X | X | X | | | | | X |
| ACTA2 | Hs00426835_g1 | | | 1 | | | | | | | | | | | | | X | | | | | | | | | |
| ANKRD22 | Hs00944015_m1 | | | 2 | | | | | | | | | | X | | | | | | | | X | | | | |
| APOL1 | Hs01066280_m1 | | | 1 | | | | | | | | | | | | | | | | | X | | | | | |
| ARG1 | Hs00968979_m1 | | | 1 | | | | | | | | X | | | | | | | | | | | | | | |
| BATF2 | Hs00912737_m1 | | | 2 | | | | | | | | | | | | | | X | X | | | | | | | |
| BCL6 | AREPURD* | | | 1 | | | | | | | | | | | | | | | | X | | | | | | |
| BLK | Hs01017452_m1 | | | 1 | | | | | | | | | | | | | | | | | | | X | | | |
| C1QA | Hs00706358_s1 | X | | 0 | | | | | | | | | | | | | | | | | | | | | | |
| C1QB | Hs00608019_m1 | | | 2 | | | | X | | | X | | | ‡ | | | | | | | | | | | | |
| C4ORF18 / FAM198B | Hs00259260_s1 | | | 1 | | | | | | | | | | X | | | | | | | | | | | | |
| C5 | Hs00156197_m1 | | | 1 | | | | | | | | | | X | | | | | | | | | | | | |
| CCR5 | Hs99999149_s1 | | | 1 | | | X | | | | | | | | | | | | | | | | | | | |
| CCR6 | Hs01890706_s1 | | | 1 | | | | | | | | | | X | | | | | | | | | | | | |
| CD160 | Hs00199894_m1 | | | 1 | | | | X | | | | | | | | | | | | | | | | | | |
| CD1C | Hs00957534_g1 | | | 1 | | | | | | | | | | | | | | | | | | | X | | | |
| CD36.2 | Hs01567186_m1 | | | 1 | | | | X | | | | | | | | | | | | | | | | | | |
| CD3E | APDJ3NT* | | | 1 | | | | | | X | | | | | | | | | | | | | | | | |
| CD40L | Hs00163934_m1 | | | 1 | | | | X | | | | | | | | | | | | | | | | | | |
| CD74-j1 | Hs04983808_s1 | X | | 0 | | | | | | | | | | | | | | | | | | | | | | |
| CD74-j2 | AP9HN47* | X | | 0 | | | | | | | | | | | | | | | | | | | | | | |
| CD79A | Hs00998120_g1 | | | 1 | | | | | | | | | | X | | | | | | | | | | | | |
| CD79B | Hs01058826_g1 | | | 1 | | | | | | | | | | X | | | | | | | | | | | | |
| CDKN1C | Hs00175938_m1 | X | | 0 | | | | | | | | | | | | | | | | | | | | | | |
| CXCR5 | Hs00540548_s1 | | | 1 | | | | | | | | | | X | | | | | | | | | | | | |
| CYP4F3 | Hs01587860_mH | | | 1 | | | | | | | | | | | | | | | | X | | | | | | |
| DUSP3 | Hs01115776_m1 | | | 2 | | | | | | | | | | X | | | | | | | | | | X | | X |
| FAM20A | Hs01034066_m1 | | | 1 | | | | | | | | | | X | | | | | | | | | | | | |
| FCGR1A | Hs00174081_m1 | | | 5 | X | | | | | | X | | X | X | | | | | | X | | | | | | |
| FCGR1A | AP2XDMK* | | | 1 | | | | | | | | | | | | | | | | | X | | | | | |
| FCGR1B | Hs02341825_m1 | | | 3 | | | | X | | | | | | X | | X | | | | | | | | | | |
| FCGR1B_ VARIANT1 | AP3267H* | | | 1 | | | | | | | | | | | | | | | | | X | | | | | |
| FCGR1B_ VARIANT2 | AP47ZTF* | | | 1 | | | | | | | | | | | | | | | | | X | | | | | |
| FCGR1C | Hs00417598_m1 | | | 1 | | | | | | | | | | X | | | | | | | | | | | | |
| FLVCR2 | Hs00900390_m1 | | | 1 | | | | | | | | | | X | | | | | | | | | | | | |
| GAS6 | AR47XGZ* | | | 2 | | | | | | | | | | X | | | | | | | | | X | | | |
| GAS6 | AR323W3* | | | 1 | | | | | | | | | | X | | | | | | | | | | | | |
| GBP1 | Hs00977005_m1 | | | 1 | | | | | | | | | | | X | | | | | | | | | | | |
| GBP2 | Hs00894846_g1 | | | 1 | | | | | | | | | | | | X | | | | | | | | | | |
| GBP5 | Hs00369472_m1 | | | 5 | X | | | | X | X | | | | | | | | | X | | | | | X | | X |
| GBP6 | Hs01584201_m1 | | | 3 | | | | X | | | | X | | ‡ | | | X | | | | | | | | | |
| GNG7 | Hs00192999_m1 | | | 1 | | | | | | | | | | X | | | | | | | | | | | | |
| GYG1 | Hs00907542_g1 | | | 1 | | | | | | | | | | | | | X | | | | | | | | | |
| GZMA | Hs00989184_m1 | | | 1 | X | | | | | | | | | | | | | | | | | | | | | |
| HK3 | Hs01092850_m1 | | | 1 | | | | | | | | | | | | | | | | X | | | | | | |
| ID3 | Hs00954037_g1 | | | 2 | | | | X | | | | | | | X | | | | | | | | | | | |
| IFI44L | Hs00915292_m1 | | | 1 | | | | | | | | | | | | | | | | X | | | | | | |
| IFITM3 | Hs03057129_s1 | | | 2 | | | | | | X | | | | | X | | | | | | | | | | | |

| Gene Symbol | TaqMan Assay ID | Assay excluded** | Reference probe | # of signatures | da Costa3 [4] | de Araujo1 [19] | Duffy4 (5)# [20] | Duffy9 (10)† [5] | Francisco2 [6] | Gjøen7 [7] | Gliddon3 [14] | Gliddon4 [14] | Jacobsen3 [8] | Kaforou22 (27)‡ [15] | Maertzdorf4 [21] | Penn-Nicholson6 [13] | Rajan5 [16] | Roe1 [22] | Roe3 [9] | Sambarey10 [10] | Satproedprai7 [11] | Suliman2 [23] | Suliman4 [23] | Sweeney3 [24,25] | Thompson5 [26] | Xpert HR [27] |
|---|---|---|---|---|---|---|---|---|---|---|---|---|---|---|---|---|---|---|---|---|---|---|---|---|---|---|
| KAZN | AP7DVDD* | | | 1 | | | | | | | | | | | | | | | | | X | | | | | |
| KIF1B | Hs01114512_g1 | | | 1 | | | | | | X | | | | | | | | | | | | | | | | |
| KLF2 | Hs00360439_g1 | | | 2 | | | | | X | | | | | | | | | | | | | | | X | | |
| KLHDC8B | Hs00293902_m1 | X | | 0 | | | | | | | | | | | | | | | | | | | | | | |
| KLRG1 | Hs00929964_m1 | | | 1 | | | X | | | | | | | | | | | | | | | | | | | |
| LAG3 | Hs00958444_g1 | | | 1 | | | | X | | | | | | | | | | | | | | | | | | |
| LHFPL2 | Hs00299613_m1 | | | 1 | | | | | | | | | | X | | | | | | | | | | | | |
| LTF | Hs00158924_m1 | | | 1 | | | | | | | | | X | | | | | | | | | | | | | |
| MAFB | APZTH2N* | | | 1 | | | | | | | | | | | | | | | | | X | | | | | |
| MAP7D3 | Hs00226257_m1 | | | 1 | | | | | | | | | | | | | | | | | | | | | X | |
| MMP9 | APEPW9P* | | | 1 | | | | | | X | | | | | | | | | | | | | | | | |
| MPO | Hs00165162_m1 | | | 1 | | | | | | | | | | X | | | | | | | | | | | | |
| MTRF1L | Hs01097882_g1 | | | 1 | | | | | | | | | | | | | X | | | | | | | | | |
| NOD2 | Hs01550759_g1 | | | 1 | | | | | | X | | | | | | | | | | | | | | | | |
| NPC2 | Hs01119244_m1 | | | 1 | | X | | | | | | | | | | | | | | | | | | | | |
| OSBPL10 | Hs00215016_m1 | | | 1 | | | | | | | | | | | | | | | | | | X | | | | |
| P2RY14 | Hs01848195_s1 | | | 1 | | | | | | | | | | | X | | | | | | | | | | | |
| PRDM1 | Hs00153357_m1 | | | 1 | | | | | | | | X | | | | | | | | | | | | | | |
| RAB13 | Hs04400188_g1 | | | 1 | | | | | | | | | | | | | | | | X | | | | | | |
| Rab33A | Hs00191243_m1 | | | 1 | | | | | | | | | X | | | | | | | | | | | | | |
| RABL2A | Hs00255244_m1 | | | 1 | | | | | | | | | | | | | X | | | | | | | | | |
| RBBP8 | Hs01090329_m1 | | | 1 | | | | | | | | | | | | | | | | X | | | | | | |
| RP11-295G20.2 | Hs01373568_m1 | | | 1 | | | | | | | | | | | | | | | | | | | | | X | |
| S100A8 | Hs00374264_g1 | | | 1 | | | | | | | | | | X | | | | | | | | | | | | |
| SCARF1 | Hs01092483_m1 | | | 1 | | | | | | | | | | | | | | | X | | | | | | | |
| SDR39U1 | Hs01016970_g1 | | | 1 | | | | | | | | | | | | X | | | | | | | | | | |
| SEPT4 | Hs00910208_g1 | | | 1 | | | | | | | | | | | | | | | | | | | X | | | |
| SERPING1 | Hs00934329_m1 | | | 1 | | | | | | | | | | | | X | | | | | | | | | | |
| SH2D1B | Hs01114628_m1 | | | 1 | | | X | | | | | | | | | | | | | | | | | | | |
| SLPI | Hs00268206_m1 | | | 1 | | | | | | | | | | | | | | | | X | | | | | | |
| SMARCD3 | Hs01088251_g1 | | | 3 | | | | | | | | | | X | | | | | | X | | | | | X | |
| STAT1 | Hs01013996_m1 | | | 1 | | | | | | | | | | | | | | | | | X | | | | | |
| STT3A | Hs00967491_m1 | | | 1 | | | | | | | | | | | | | | | | | | | | | X | |
| TIMM10 | APT2DEC* | | | 1 | | | | | | | | | | | | | | | | X | | | | | | |
| TMCC1 | Hs01037666_s1 | | | 1 | | | | | | | | X | | | | | | | | | | | | | | |
| TNIP1 | AP47ZYZ* | | | 1 | | | | | | X | | | | | | | | | | | | | | | | |
| TPPP3 | Hs00372228_g1 | | | 1 | | | X | | | | | | | | | | | | | | | | | | | |
| TRMT2A | Hs01000041_g1 | | | 1 | | | | | | | | | | | | X | | | | | | | | | | |
| TUBGCP6 | Hs00363509_g1 | | | 1 | | | | | | | | | | | | X | | | | | | | | | | |
| UCP2 | Hs01075224_g1 | | | 1 | | | | | | | | | | | | | | | | | | | | | X | |
| VAMP5 | Hs01105383_g1 | | | 1 | | | | | | | | | | X | | | | | | | | | | | | |
| WARS-j1 | Hs00998737_m1 | X | | 0 | | | | | | | | | | | | | | | | | | | | | | |
| WARS-j2 | APAAFHX* | X | | 0 | | | | | | | | | | | | | | | | | | | | | | |
| ZDHHC19 | Hs00376118_m1 | | | 1 | | | | X | | | | | | | | | | | | | | | | | | |
| ZFYVE9 | Hs01024382_m1 | | | 1 | | | # | | | | | | | | | | | | | | | | | | | |
| ZNF296 | Hs00377132_m1 | | | 2 | | | | | | | X | | | X | | | | | | | | | | | | |

Signatures are named by first author and number of transcripts included in the model (e.g. Author11). Numbers in brackets indicate the original number of transcripts in the published model. Some signatures have a reduced number of transcripts when translated to RT-qPCR due to duplicate transcript symbols (IDs), high-primer probe failure rate (**poor amplification efficiency), or where transcript sequences from an original discovery cohorts could not be mapped to a more recent reference transcriptome.
* Custom designed primer-probe assays; available on request to corresponding author.
### One transcripts (ZFYVE9) was excluded from the Duffy5 signature due to a high primer-probe assay failure rate.
† The CERKL1 primer-probe assay failed during panel optimisation and was removed from the Duffy10 signature.
‡ Three transcripts (C1QB, C1QC, and GBP6) were excluded from the Kaforou27 signature due to a high primer-probe assay failure rate. Unique primer-probe assays could not be designed for both of the FCGR1B transcript variants; only one FCGR1B primer-probe was included in the Kaforou22 model. We were unable to design a primer-probe for LOC728744; all information had been withdrawn from both the NCBI and RefSeq databases.
Xpert HR=Xpert Host-Response.

**Table S2.** Transcriptomic signatures on the panel that were reparameterised.

| Signature | Reference | Original gene expression measurement | Model used | HIV-negative training and test set (combined) AUC (95% CI) | HIV-positive validation set AUC (95% CI) |
|---|---|---|---|---|---|
| **Da Costa3** | *Tuberculosis* (2015)[4] | RT-qPCR | Random forest | 0.98 (0.93−1) | 0.96 (0.93−1) |
| **Duffy9** | *PLoS One* (2019)[5] | Meta-analysis of microarray data | Multinomial random forest | TB vs LTBI: 1 (1−1)* | |
| **Francisco2** | *J Infect* (2017)[6] | RT-qPCR | Random forest | 0.99 (0.98−1) | 0.83 (0.74−0.93) |
| **Gjøen7** | *Sci Rep* (2017)[7] | Dual-color-Reverse-Transcriptase-Multiplex-Ligation-dependent-Probe-Amplification (dc-RT MLPA) | LASSO regression | 0.98 (0.94−1) | 0.78 (0.67−0.88) |
| **Jacobsen3** | *J Mol Med* (2007)[8] | RT-qPCR | Linear discriminant analysis | 0.99 (0.99−1) | 0.95 (0.89−1) |
| **Roe3** | *Clin Infect Dis* (2020)[9] | RNA sequencing | Support vector machines (linear kernel) | 0.96 (0.89−1) | 0.89 (0.80−0.97) |
| **Sambarey10** | *EBioMedicine* (2017)[10] | RNA sequencing | Linear discriminant analysis | 0.99 (0.96−1) | 0.95 (0.90−1) |
| **Satproedprai7** | *Genes Immun* (2015)[11] | RT-qPCR | LASSO regression | 0.99 (0.98−1) | 0.95 (0.90 −1) |

* HIV-positive samples required to parameterise the multinomial model. No independent validation set available.

**Table S3.** Target Product Profile for TB treatment biomarkers.

| Timing of test | Use Case | Notes | Sensitivity | | Specificity | |
|---|---|---|---|---|---|---|
| | | | **Minimum** | **Optimum** | **Minimum** | **Optimum** |
| **Baseline/ diagnosis**<br>**Treatment initiation** | Identify those who require more intensive treatment regimen OR shorter regimen | To avoid undertreatment of those with higher risk<br>(e.g., more severe disease) | **>90%** | >95% | **>70%** | >80% |
| **During treatment** | Identify those at risk of poor outcome OR for shorter regimen | Identify those who do not adequately respond<br>Adherence support | **>75%** | >90% | **>80%** | >90% |
| **End of treatment** | Identify those with poor outcome | Prolong treatment | **>80%** | >95% | **>90%** | >95% |

Reported by Gupta-Wright et al. (2023)[28] and WHO (2023)[29]

**Table S4.** Performance (area under the curve) of transcriptomic signatures measured at baseline, month 2, and end of treatment (month 6) for predicting TB recurrence after completion of treatment.

| Signature | Baseline controls | Baseline cases | Baseline AUC (95% CI) | Month 2 controls | Month 2 cases | Month 2 AUC (95% CI) | End of treatment controls | End of treatment cases | End of treatment AUC (95% CI) |
|---|---|---|---|---|---|---|---|---|---|
| **da Costa3** | 259 | 9 | 0.71 (0.60–0.82) | 261 | 9 | 0.80 (0.67–0.94) | 259 | 9 | 0.84 (0.70–0.98) |
| **de Araujo1** | 259 | 9 | 0.91 (0.83–0.98) | 261 | 9 | 0.88 (0.79–0.98) | 259 | 9 | 0.77 (0.62–0.92) |
| **Duffy4** | 246 | 9 | 0.83 (0.67–0.99) | 257 | 8 | 0.77 (0.60–0.93) | 256 | 9 | 0.64 (0.47–0.80) |
| **Duffy9** | 249 | 9 | 0.78 (0.63–0.94) | 247 | 8 | 0.85 (0.71–0.98) | 236 | 9 | 0.73 (0.57–0.89) |
| **Francisco2** | 248 | 7 | 0.80 (0.72–0.89) | 252 | 7 | 0.78 (0.54–1.00) | 248 | 9 | 0.82 (0.66–0.98) |
| **Gjoen7** | 259 | 9 | 0.82 (0.71–0.92) | 258 | 9 | 0.84 (0.68–1.00) | 259 | 9 | 0.89 (0.82–0.97) |
| **Gliddon3** | 243 | 6 | 0.91 (0.83–0.99) | 243 | 6 | 0.84 (0.72–0.97) | 234 | 8 | 0.71 (0.49–0.93) |
| **Gliddon4** | 254 | 9 | 0.85 (0.76–0.95) | 240 | 9 | 0.78 (0.58–0.98) | 220 | 9 | 0.59 (0.35–0.83) |
| **Jacobsen3** | 217 | 7 | 0.75 (0.62–0.87) | 209 | 7 | 0.81 (0.64–0.98) | 200 | 9 | 0.81 (0.64–0.99) |
| **Kaforou22** | 227 | 6 | 0.91 (0.83–0.99) | 215 | 6 | 0.87 (0.71–1.00) | 182 | 6 | 0.75 (0.49–1.00) |
| **Maertzdorf4** | 251 | 8 | 0.82 (0.64–0.99) | 254 | 8 | 0.80 (0.58–1.00) | 250 | 9 | 0.76 (0.55–0.97) |
| **Penn-Nicholson6** | 258 | 9 | 0.78 (0.60–0.96) | 260 | 8 | 0.79 (0.61–0.97) | 258 | 9 | 0.77 (0.58–0.97) |
| **Rajan5** | 215 | 7 | 0.72 (0.57–0.88) | 193 | 8 | 0.73 (0.53–0.93) | 175 | 9 | 0.67 (0.49–0.85) |
| **Roe1** | 258 | 9 | 0.78 (0.68–0.88) | 258 | 9 | 0.82 (0.69–0.95) | 255 | 9 | 0.73 (0.52–0.94) |
| **Roe3** | 257 | 9 | 0.85 (0.74–0.95) | 258 | 9 | 0.82 (0.66–0.99) | 251 | 9 | 0.83 (0.70–0.95) |
| **Sambarey10** | 254 | 9 | 0.76 (0.67–0.86) | 255 | 8 | 0.82 (0.67–0.98) | 254 | 9 | 0.77 (0.57–0.97) |
| **Satproedprai7** | 254 | 9 | 0.88 (0.79–0.97) | 249 | 9 | 0.86 (0.72–1.00) | 244 | 9 | 0.82 (0.65–0.99) |
| **Suliman2** | 149 | 2 | 0.76 (0.35–1.00) | 150 | 4 | 0.80 (0.61–1.00) | 163 | 8 | 0.67 (0.47–0.87) |
| **Suliman4** | 249 | 9 | 0.71 (0.51–0.90) | 252 | 8 | 0.84 (0.78–0.91) | 247 | 9 | 0.64 (0.47–0.82) |
| **Sweeney3** | 248 | 7 | 0.82 (0.70–0.94) | 252 | 7 | 0.78 (0.59–0.97) | 246 | 9 | 0.73 (0.54–0.93) |
| **Thompson5** | 257 | 9 | 0.79 (0.68–0.89) | 261 | 8 | 0.79 (0.65–0.94) | 258 | 9 | 0.42 (0.19–0.65) |
| **Xpert HR** | 259 | 9 | 0.80 (0.66–0.94) | 261 | 9 | 0.84 (0.72–0.95) | 257 | 9 | 0.84 (0.71–0.96) |

AUC=area under the curve. CI=confidence interval. Xpert HR=Xpert Host-Response.

**Table S5.** Sensitivity, specificity, positive predictive value, and negative predictive value of transcriptomic signatures measured at baseline, month 2, and end of treatment (month 6) for predicting TB recurrence after completion of treatment.

| Signature | Baseline sensitivity, % (95% CI) | Baseline specificity, % (95% CI) | Month 2 sensitivity, % (95% CI) | Month 2 specificity, % (95% CI) | End of treatment sensitivity, % (95% CI) | End of treatment specificity, % (95% CI) |
|---|---|---|---|---|---|---|
| **WHO Target Product Profile for TB treatment biomarkers** | Minimum: 90%<br>Optimum: 95% | **Minimum: 70% ***<br>Optimum: 80% | Minimum: 75%<br>Optimum: 90% | **Minimum: 80% ***<br>Optimum: 90% | Minimum: 80%<br>Optimum: 95% | **Minimum: 90% ***<br>Optimum: 95% |
| **da Costa3** | 55.6 (26.7– 81.1) | 69.9 (64.0–75.1) | 55.6 (26.7– 81.1) | 79.7 (74.4–84.1) | 55.6 (26.7– 81.1) | 89.6 (85.3–92.7) |
| **de Araujo1** | 88.9 (56.5– 98.0) | 69.9 (64.0–75.1) | 88.9 (56.5– 98.0) | 79.7 (74.4–84.1) | 33.3 (12.1– 64.6) | 89.6 (85.3–92.7) |
| **Duffy4** | 88.9 (56.5– 98.0) | 69.5 (63.5–74.9) | 62.5 (30.6– 86.3) | 79.8 (74.4–84.2) | 22.2 ( 6.3– 54.7) | 89.8 (85.5–93.0) |
| **Duffy9** | 77.8 (45.3– 93.7) | 69.9 (63.9–75.2) | 87.5 (52.9– 97.8) | 79.8 (74.3–84.3) | 33.3 (12.1– 64.6) | 89.8 (85.3–93.1) |
| **Francisco2** | 85.7 (48.7– 97.4) | 70.6 (64.6–75.9) | 71.4 (35.9– 91.8) | 80.6 (75.2–85.0) | 66.7 (35.4– 87.9) | 89.5 (85.1–92.7) |
| **Gjoen7** | 88.9 (56.5– 98.0) | 69.9 (64.0–75.1) | 77.8 (45.3– 93.7) | 79.8 (74.5–84.3) | 66.7 (35.4– 87.9) | 89.6 (85.3–92.7) |
| **Gliddon3** | 100.0 (61.0–100.0) | 69.5 (63.5–75.0) | 66.7 (30.0– 90.3) | 79.8 (74.3–84.4) | 37.5 (13.7– 69.4) | 89.7 (85.2–93.0) |
| **Gliddon4** | 77.8 (45.3– 93.7) | 69.7 (63.8–75.0) | 77.8 (45.3– 93.7) | 79.6 (74.0–84.2) | 33.3 (12.1– 64.6) | 89.5 (84.8–92.9) |
| **Jacobsen3** | 42.9 (15.8– 75.0) | 69.6 (63.2–75.3) | 71.4 (35.9– 91.8) | 79.9 (74.0–84.8) | 44.4 (18.9– 73.3) | 89.5 (84.5–93.0) |
| **Kaforou22** | 100.0 (61.0–100.0) | 69.6 (63.3–75.2) | 83.3 (43.6– 97.0) | 79.5 (73.6–84.4) | 50.0 (18.8– 81.2) | 89.6 (84.3–93.2) |
| **Maertzdorf4** | 75.0 (40.9– 92.9) | 69.7 (63.8–75.1) | 87.5 (52.9– 97.8) | 79.5 (74.1–84.0) | 44.4 (18.9– 73.3) | 89.6 (85.2–92.8) |
| **Penn-Nicholson6** | 77.8 (45.3– 93.7) | 69.8 (63.9–75.0) | 75.0 (40.9– 92.9) | 79.6 (74.3–84.1) | 44.4 (18.9– 73.3) | 89.5 (85.2–92.7) |
| **Rajan5** | 57.1 (25.0– 84.2) | 69.8 (63.3–75.5) | 50.0 (21.5– 78.5) | 79.8 (73.6–84.9) | 22.2 ( 6.3– 54.7) | 89.7 (84.3–93.4) |
| **Roe1** | 77.8 (45.3– 93.7) | 69.8 (63.9–75.0) | 55.6 (26.7– 81.1) | 79.8 (74.5–84.3) | 44.4 (18.9– 73.3) | 89.8 (85.5–92.9) |
| **Roe3** | 77.8 (45.3– 93.7) | 69.6 (63.8–74.9) | 77.8 (45.3– 93.7) | 79.8 (74.5–84.3) | 55.6 (26.7– 81.1) | 89.6 (85.3–92.8) |
| **Sambarey10** | 66.7 (35.4– 87.9) | 69.7 (63.8–75.0) | 75.0 (40.9– 92.9) | 79.6 (74.2–84.1) | 55.6 (26.7– 81.1) | 89.8 (85.4–92.9) |
| **Satproedprai7** | 88.9 (56.5– 98.0) | 69.7 (63.8–75.0) | 88.9 (56.5– 98.0) | 79.5 (74.1–84.1) | 44.4 (18.9– 73.3) | 89.8 (85.3–93.0) |
| **Suliman2** | 50.0 ( 9.5– 90.5) | 69.8 (62.0–76.6) | 75.0 (30.1– 95.4) | 80.0 (72.9–85.6) | 12.5 ( 2.2– 47.1) | 89.6 (83.9–93.4) |
| **Suliman4** | 66.7 (35.4– 87.9) | 69.9 (63.9–75.2) | 75.0 (40.9– 92.9) | 79.8 (74.4–84.3) | 11.1 ( 2.0– 43.5) | 89.9 (85.5–93.0) |
| **Sweeney3** | 71.4 (35.9– 91.8) | 69.8 (63.8–75.1) | 71.4 (35.9– 91.8) | 79.8 (74.4–84.3) | 22.2 ( 6.3– 54.7) | 89.8 (85.4–93.0) |
| **Thompson5** | 66.7 (35.4– 87.9) | 69.6 (63.8–74.9) | 62.5 (30.6– 86.3) | 79.7 (74.4–84.1) | 11.1 ( 2.0– 43.5) | 89.5 (85.2–92.7) |
| **Xpert HR** | 77.8 (45.3– 93.7) | 69.9 (64.0–75.1) | 88.9 (56.5– 98.0) | 79.7 (74.4–84.1) | 55.6 (26.7– 81.1) | 89.9 (85.6–93.0) |

* Signatures benchmarked at 70%, 80%, and 90% specificity at baseline, month 2, and end of treatment, respectively.

CI=confidence interval. Xpert HR=Xpert Host-Response.

**Table S6.** Performance (area under the curve) of transcriptomic signatures measured at baseline, month 2, and end of treatment (month 6) for predicting TB treatment failure.

| Signature | Baseline controls | Baseline cases | Baseline AUC (95% CI) | Month 2 controls | Month 2 cases | Month 2 AUC (95% CI) | End of treatment controls | End of treatment cases | End of treatment AUC (95% CI) |
|---|---|---|---|---|---|---|---|---|---|
| **da Costa3** | 259 | 32 | 0.64 (0.54–0.73) | 261 | 31 | 0.55 (0.45–0.65) | 259 | 28 | 0.60 (0.48–0.71) |
| **de Araujo1** | 259 | 32 | 0.69 (0.60–0.77) | 261 | 31 | 0.56 (0.46–0.66) | 259 | 28 | 0.62 (0.51–0.73) |
| **Duffy4** | 246 | 28 | 0.62 (0.51–0.72) | 257 | 29 | 0.63 (0.52–0.73) | 256 | 27 | 0.57 (0.46–0.68) |
| **Duffy9** | 249 | 31 | 0.57 (0.46–0.69) | 247 | 28 | 0.53 (0.41–0.64) | 236 | 24 | 0.62 (0.51–0.72) |
| **Francisco2** | 248 | 26 | 0.68 (0.57–0.80) | 252 | 28 | 0.51 (0.40–0.62) | 248 | 26 | 0.58 (0.46–0.70) |
| **Gjoen7** | 259 | 32 | 0.66 (0.56–0.76) | 258 | 31 | 0.53 (0.42–0.63) | 259 | 28 | 0.47 (0.36–0.58) |
| **Gliddon3** | 243 | 26 | 0.56 (0.44–0.68) | 243 | 29 | 0.55 (0.43–0.67) | 234 | 25 | 0.45 (0.33–0.57) |
| **Gliddon4** | 254 | 31 | 0.58 (0.47–0.69) | 240 | 29 | 0.53 (0.42–0.63) | 220 | 22 | 0.52 (0.37–0.68) |
| **Jacobsen3** | 217 | 26 | 0.61 (0.50–0.72) | 209 | 29 | 0.57 (0.46–0.68) | 200 | 26 | 0.59 (0.47–0.72) |
| **Kaforou22** | 227 | 22 | 0.49 (0.38–0.61) | 215 | 24 | 0.54 (0.41–0.67) | 182 | 16 | 0.60 (0.41–0.78) |
| **Maertzdorf4** | 251 | 32 | 0.58 (0.47–0.68) | 254 | 29 | 0.49 (0.38–0.60) | 250 | 27 | 0.53 (0.43–0.64) |
| **Penn-Nicholson6** | 258 | 32 | 0.63 (0.53–0.74) | 260 | 31 | 0.55 (0.45–0.65) | 258 | 28 | 0.59 (0.49–0.70) |
| **Rajan5** | 215 | 26 | 0.51 (0.38–0.63) | 193 | 28 | 0.62 (0.50–0.74) | 175 | 22 | 0.50 (0.35–0.65) |
| **Roe1** | 258 | 32 | 0.62 (0.53–0.71) | 258 | 31 | 0.52 (0.42–0.63) | 255 | 26 | 0.57 (0.45–0.69) |
| **Roe3** | 257 | 32 | 0.69 (0.59–0.78) | 258 | 31 | 0.50 (0.40–0.60) | 251 | 26 | 0.52 (0.39–0.64) |
| **Sambarey10** | 254 | 31 | 0.64 (0.54–0.74) | 255 | 31 | 0.55 (0.45–0.66) | 254 | 27 | 0.61 (0.49–0.73) |
| **Satproedprai7** | 254 | 32 | 0.64 (0.54–0.73) | 249 | 28 | 0.59 (0.48–0.70) | 244 | 27 | 0.62 (0.50–0.73) |
| **Suliman2** | 149 | 17 | 0.55 (0.41–0.69) | 150 | 18 | 0.52 (0.37–0.66) | 163 | 20 | 0.65 (0.53–0.77) |
| **Suliman4** | 249 | 31 | 0.50 (0.39–0.61) | 252 | 30 | 0.51 (0.40–0.62) | 247 | 26 | 0.53 (0.41–0.66) |
| **Sweeney3** | 248 | 26 | 0.66 (0.55–0.76) | 252 | 28 | 0.52 (0.41–0.62) | 246 | 26 | 0.53 (0.41–0.64) |
| **Thompson5** | 257 | 31 | 0.66 (0.56–0.76) | 261 | 31 | 0.63 (0.53–0.73) | 258 | 28 | 0.66 (0.56–0.77) |
| **Xpert HR** | 259 | 32 | 0.57 (0.47–0.66) | 261 | 31 | 0.50 (0.40–0.61) | 257 | 28 | 0.52 (0.39–0.65) |

AUC=area under the curve. CI=confidence interval. Xpert HR=Xpert Host-Response.

**Table S7.** Sensitivity, specificity, positive predictive value, and negative predictive value of transcriptomic signatures measured at baseline, month 2, and end of treatment (month 6) for predicting TB treatment failure.

| Signature | Baseline sensitivity, % (95% CI) | Baseline specificity, % (95% CI) | Month 2 sensitivity, % (95% CI) | Month 2 specificity, % (95% CI) | End of treatment sensitivity, % (95% CI) | End of treatment specificity, % (95% CI) |
|---|---|---|---|---|---|---|
| **WHO Target Product Profile for TB treatment biomarkers** | Minimum: 90%<br>Optimum: 95% | **Minimum: 70% ***<br>Optimum: 80% | Minimum: 75%<br>Optimum: 90% | **Minimum: 80% ***<br>Optimum: 90% | Minimum: 80%<br>Optimum: 95% | **Minimum: 90% ***<br>Optimum: 95% |
| **da Costa3** | 40.6 (25.5–57.7) | 69.9 (64.0–75.1) | 19.4 ( 9.2–36.3) | 79.7 (74.4–84.1) | 17.9 ( 7.9–35.6) | 89.6 (85.3–92.7) |
| **de Araujo1** | 50.0 (33.6–66.4) | 69.9 (64.0–75.1) | 12.9 ( 5.1–28.9) | 79.7 (74.4–84.1) | 14.3 ( 5.7–31.5) | 89.6 (85.3–92.7) |
| **Duffy4** | 53.6 (35.8–70.5) | 69.5 (63.5–74.9) | 37.9 (22.7–56.0) | 79.8 (74.4–84.2) | 14.8 ( 5.9–32.5) | 89.8 (85.5–93.0) |
| **Duffy9** | 45.2 (29.2–62.2) | 69.9 (63.9–75.2) | 25.0 (12.7–43.4) | 79.8 (74.3–84.3) | 8.3 ( 2.3–25.8) | 89.8 (85.3–93.1) |
| **Francisco2** | 57.7 (38.9–74.5) | 70.6 (64.6–75.9) | 14.3 ( 5.7–31.5) | 80.6 (75.2–85.0) | 23.1 (11.0–42.1) | 89.5 (85.1–92.7) |
| **Gjoen7** | 53.1 (36.4–69.1) | 69.9 (64.0–75.1) | 19.4 ( 9.2–36.3) | 79.8 (74.5–84.3) | 3.6 ( 0.6–17.7) | 89.6 (85.3–92.7) |
| **Gliddon3** | 38.5 (22.4–57.5) | 69.5 (63.5–75.0) | 20.7 ( 9.8–38.4) | 79.8 (74.3–84.4) | 8.0 ( 2.2–25.0) | 89.7 (85.2–93.0) |
| **Gliddon4** | 38.7 (23.7–56.2) | 69.7 (63.8–75.0) | 20.7 ( 9.8–38.4) | 79.6 (74.0–84.2) | 22.7 (10.1–43.4) | 89.5 (84.8–92.9) |
| **Jacobsen3** | 42.3 (25.5–61.1) | 69.6 (63.2–75.3) | 27.6 (14.7–45.7) | 79.9 (74.0–84.8) | 23.1 (11.0–42.1) | 89.5 (84.5–93.0) |
| **Kaforou22** | 18.2 ( 7.3–38.5) | 69.6 (63.3–75.2) | 25.0 (12.0–44.9) | 79.5 (73.6–84.4) | 31.2 (14.2–55.6) | 89.6 (84.3–93.2) |
| **Maertzdorf4** | 46.9 (30.9–63.6) | 69.7 (63.8–75.1) | 24.1 (12.2–42.1) | 79.5 (74.1–84.0) | 7.4 ( 2.1–23.4) | 89.6 (85.2–92.8) |
| **Penn-Nicholson6** | 50.0 (33.6–66.4) | 69.8 (63.9–75.0) | 19.4 ( 9.2–36.3) | 79.6 (74.3–84.1) | 17.9 ( 7.9–35.6) | 89.5 (85.2–92.7) |
| **Rajan5** | 30.8 (16.5–50.0) | 69.8 (63.3–75.5) | 35.7 (20.7–54.2) | 79.8 (73.6–84.9) | 13.6 ( 4.7–33.3) | 89.7 (84.3–93.4) |
| **Roe1** | 40.6 (25.5–57.7) | 69.8 (63.9–75.0) | 19.4 ( 9.2–36.3) | 79.8 (74.5–84.3) | 15.4 ( 6.2–33.5) | 89.8 (85.5–92.9) |
| **Roe3** | 50.0 (33.6–66.4) | 69.6 (63.8–74.9) | 12.9 ( 5.1–28.9) | 79.8 (74.5–84.3) | 11.5 ( 4.0–29.0) | 89.6 (85.3–92.8) |
| **Sambarey10** | 48.4 (32.0–65.2) | 69.7 (63.8–75.0) | 29.0 (16.1–46.6) | 79.6 (74.2–84.1) | 22.2 (10.6–40.8) | 89.8 (85.4–92.9) |
| **Satproedprai7** | 43.8 (28.2–60.7) | 69.7 (63.8–75.0) | 25.0 (12.7–43.4) | 79.5 (74.1–84.1) | 18.5 ( 8.2–36.7) | 89.8 (85.3–93.0) |
| **Suliman2** | 35.3 (17.3–58.7) | 69.8 (62.0–76.6) | 27.8 (12.5–50.9) | 80.0 (72.9–85.6) | 10.0 ( 2.8–30.1) | 89.6 (83.9–93.4) |
| **Suliman4** | 41.9 (26.4–59.2) | 69.9 (63.9–75.2) | 16.7 ( 7.3–33.6) | 79.8 (74.4–84.3) | 19.2 ( 8.5–37.9) | 89.9 (85.5–93.0) |
| **Sweeney3** | 46.2 (28.8–64.5) | 69.8 (63.8–75.1) | 17.9 ( 7.9–35.6) | 79.8 (74.4–84.3) | 7.7 ( 2.1–24.1) | 89.8 (85.4–93.0) |
| **Thompson5** | 58.1 (40.8–73.6) | 69.6 (63.8–74.9) | 25.8 (13.7–43.2) | 79.7 (74.4–84.1) | 17.9 ( 7.9–35.6) | 89.5 (85.2–92.7) |
| **Xpert HR** | 25.0 (13.3–42.1) | 69.9 (64.0–75.1) | 16.1 ( 7.1–32.6) | 79.7 (74.4–84.1) | 17.9 ( 7.9–35.6) | 89.9 (85.6–93.0) |

* Signatures benchmarked at 70%, 80%, and 90% specificity at baseline, month 2, and end of treatment, respectively.

CI=confidence interval. Xpert HR=Xpert Host-Response.

**Table S8.** Performance (area under the curve) of transcriptomic signatures measured at baseline for predicting death due to TB or unknown cause.

| Signature | Baseline controls | Baseline cases | Baseline AUC (95% CI) |
|---|---|---|---|
| **da Costa3** | 259 | 20 | 0.52 (0.39–0.65) |
| **de Araujo1** | 259 | 20 | 0.54 (0.39–0.68) |
| **Duffy4** | 246 | 19 | 0.81 (0.70–0.92) |
| **Duffy9** | 249 | 19 | 0.72 (0.62–0.81) |
| **Francisco2** | 248 | 20 | 0.56 (0.42–0.70) |
| **Gjoen7** | 259 | 20 | 0.54 (0.39–0.68) |
| **Gliddon3** | 243 | 17 | 0.71 (0.61–0.81) |
| **Gliddon4** | 254 | 18 | 0.76 (0.64–0.87) |
| **Jacobsen3** | 217 | 13 | 0.73 (0.64–0.82) |
| **Kaforou22** | 227 | 16 | 0.73 (0.62–0.85) |
| **Maertzdorf4** | 251 | 20 | 0.73 (0.63–0.84) |
| **Penn-Nicholson6** | 258 | 20 | 0.69 (0.59–0.78) |
| **Rajan5** | 215 | 12 | 0.54 (0.35–0.73) |
| **Roe1** | 258 | 20 | 0.53 (0.41–0.65) |
| **Roe3** | 257 | 20 | 0.53 (0.37–0.68) |
| **Sambarey10** | 254 | 20 | 0.69 (0.61–0.77) |
| **Satproedprai7** | 254 | 20 | 0.70 (0.60–0.81) |
| **Suliman2** | 149 | 4 | 0.54 (0.29–0.79) |
| **Suliman4** | 249 | 20 | 0.62 (0.49–0.75) |
| **Sweeney3** | 248 | 20 | 0.69 (0.55–0.82) |
| **Thompson5** | 257 | 19 | 0.82 (0.73–0.90) |
| **Xpert HR** | 259 | 20 | 0.52 (0.37–0.67) |

AUC=area under the curve. CI=confidence interval. Xpert HR=Xpert Host-Response.

**Table S9.** Sensitivity, specificity, positive predictive value, and negative predictive value of transcriptomic signatures measured at baseline for predicting death due to TB or unknown cause.

| Signature | Baseline sensitivity, % (95% CI) | Baseline specificity, % (95% CI) |
|---|---|---|
| **WHO Target Product Profile for TB treatment biomarkers** | Minimum: 90%<br>Optimum: 95% | **Minimum: 70% ***<br>Optimum: 80% |
| **da Costa3** | 40.0 (21.9–61.3) | 69.9 (64.0–75.1) |
| **de Araujo1** | 45.0 (25.8–65.8) | 69.9 (64.0–75.1) |
| **Duffy4** | 78.9 (56.7–91.5) | 69.5 (63.5–74.9) |
| **Duffy9** | 57.9 (36.3–76.9) | 69.9 (63.9–75.2) |
| **Francisco2** | 35.0 (18.1–56.7) | 70.6 (64.6–75.9) |
| **Gjoen7** | 40.0 (21.9–61.3) | 69.9 (64.0–75.1) |
| **Gliddon3** | 52.9 (31.0–73.8) | 69.5 (63.5–75.0) |
| **Gliddon4** | 66.7 (43.7–83.7) | 69.7 (63.8–75.0) |
| **Jacobsen3** | 53.8 (29.1–76.8) | 69.6 (63.2–75.3) |
| **Kaforou22** | 62.5 (38.6–81.5) | 69.6 (63.3–75.2) |
| **Maertzdorf4** | 65.0 (43.3–81.9) | 69.7 (63.8–75.1) |
| **Penn-Nicholson6** | 40.0 (21.9–61.3) | 69.8 (63.9–75.0) |
| **Rajan5** | 41.7 (19.3–68.0) | 69.8 (63.3–75.5) |
| **Roe1** | 30.0 (14.5–51.9) | 69.8 (63.9–75.0) |
| **Roe3** | 45.0 (25.8–65.8) | 69.6 (63.8–74.9) |
| **Sambarey10** | 35.0 (18.1–56.7) | 69.7 (63.8–75.0) |
| **Satproedprai7** | 60.0 (38.7–78.1) | 69.7 (63.8–75.0) |
| **Suliman2** | 25.0 ( 4.6–69.9) | 69.8 (62.0–76.6) |
| **Suliman4** | 40.0 (21.9–61.3) | 69.9 (63.9–75.2) |
| **Sweeney3** | 70.0 (48.1–85.5) | 69.8 (63.8–75.1) |
| **Thompson5** | 84.2 (62.4–94.5) | 69.6 (63.8–74.9) |
| **Xpert HR** | 45.0 (25.8–65.8) | 69.9 (64.0–75.1) |

* Signatures benchmarked at 70% specificity.

CI=confidence interval. Xpert HR=Xpert Host-Response.

**Table S10.** Multivariable linear regression analysis to identify factors associated with baseline Thompson5 and XpertHR signature scores.

| | Thompson5 | | Xpert HR | |
|---|---|---|---|---|
| | **β coefficient (95% CI)** | **p-value** | **β coefficient (95% CI)** | **p-value** |
| (Intercept) | 0.75 (0.52; 0.97) | <0.001 | 0.83 (0.59; 1.07) | <0.001 |
| Sex | | | | |
| Female | Reference | | Reference | |
| Male | 0.04 (-0.03; 0.10) | 0.25 | 0.01 (-0.05; 0.07) | 0.76 |
| Age (per 10 years) | -0.01 (-0.03; 0.01) | 0.39 | -0.03 (-0.05; -0.01) | **0.003** |
| City | | | | |
| Manaus | Reference | | Reference | |
| Rio de Janeiro | -0.06 (-0.14; 0.03) | 0.22 | 0 (-0.09; 0.09) | >0.99 |
| Salvador | 0 (-0.09; 0.10) | 0.98 | 0.03 (-0.07; 0.13) | 0.49 |
| Ethnicity | | | | |
| Black | Reference | | Reference | |
| Pardo | 0.05 (-0.04; 0.13) | 0.26 | -0.02 (-0.11; 0.07) | 0.63 |
| Indian | 0.17 (-0.05; 0.38) | 0.13 | 0.12 (-0.11; 0.35) | 0.31 |
| White | 0.04 (-0.05; 0.14) | 0.37 | -0.08 (-0.19; 0.02) | 0.11 |
| Unknown | -0.12 (-0.57; 0.33) | 0.60 | -0.13 (-0.60; 0.34) | 0.59 |
| Smoking history | | | | |
| Never smoked | Reference | | Reference | |
| Former smoker | -0.01 (-0.06; 0.04) | 0.64 | -0.01 (-0.06; 0.04) | 0.57 |
| Current smoker | 0.05 (-0.01; 0.11) | 0.13 | 0.04 (-0.03; 0.10) | 0.28 |
| Baseline body-mass index (per 10 kg/m$^2$) | -0.01 (-0.02; 0) | 0.087 | 0 (-0.01; 0.010) | 0.45 |
| Baseline HIV status | | | | |
| Negative | Reference | | Reference | |
| Positive | -0.04 (-0.12; 0.03) | 0.25 | -0.04 (-0.12; 0.04) | 0.33 |
| Sputum smear status | | | | |
| Scanty | Reference | | Reference | |
| 1+ | 0.06 (-0.03; 0.15) | 0.19 | 0.07 (-0.02; 0.17) | 0.13 |
| 2+ | 0.05 (-0.05; 0.14) | 0.35 | **0.11 (0.01; 0.21)** | **0.038** |
| 3+ | 0.10 (0; 0.20) | **0.040** | **0.15 (0.05; 0.25)** | **0.004** |
| Chest radiographic evidence of cavitation | | | | |
| No | Reference | | Reference | |
| Yes | 0.04 (-0.03; 0.11) | 0.26 | -0.04 (-0.11; 0.03) | 0.30 |
| Uncertain | 0.05 (-0.06; 0.16) | 0.40 | 0 (-0.12; 0.11) | 0.99 |
| TB disease outcome | | | | |
| Recurrence-free cure | Reference | | Reference | |
| Death due to TB or unknown causes | **0.18 (0.07; 0.29)** | **0.002** | 0.04 (-0.08; 0.16) | 0.51 |
| Treatment failure | **0.09 (0; 0.19)** | **0.045** | 0.04 (-0.06; 0.13) | 0.43 |
| TB recurrence | **0.23 (0.06; 0.40)** | **0.007** | **0.23 (0.05; 0.40)** | **0.014** |
| **Summary Statistics** | | | | |
| Adjusted $R^2$ | 0.12 | | 0.07 | |
| Missing variables | 72 | | 73 | |
| Degrees of freedom | 227 | | 230 | |

CI=confidence interval. Xpert HR=Xpert Host-Response
